# EpiSignaLR: an open resource for genome-wide episignature analysis from methylation-aware short- and long-read sequencing

**DOI:** 10.64898/2026.10.21.26366070

**Authors:** Sophia B. Gibson, Miranda P.G. Zalusky, Joy Goffena, Elizabeth J. Hamlin, Ruby Lamb, Jenna R. Rosinski, Trent Prall, Zachary B. Anderson, Sophie H.R. Storz, Angela L. Miller, Yuwei Shi, Kristina Lanko, David Mulder, Yaoqing Shen, Kim Lensing, Martin G. Elferink, Samantha Vergano, Monica H. Wojcik, Koen L.I. van Gassen, William T. Gibson, Steven J.M. Jones, Tahsin Stefan Barakat, Danny E. Miller

**Author notes:** Correspondence: Danny E. Miller.

## Abstract

**Background:** Episignatures, disorder-specific patterns of genome-wide DNA methylation, are established for more than 100 Mendelian conditions and help resolve variants of uncertain significance and evaluate unsolved cases. Current clinical testing approaches typically use data generated by methylation arrays, which require a specific assay and limit discovery to the probe set of the array. Long-read and Illumina 5-base genome sequencing measure CpG methylation alongside genetic variation, raising the possibility of folding episignature analysis into a single genome-wide test.

**Methods:** We sequenced individuals with *SETD1B*-related neurodevelopmental disorder, Kabuki syndrome, or Kleefstra syndrome, along with controls, using Oxford Nanopore Technologies (ONT), PacBio HiFi, or Illumina 5-base whole-genome sequencing. We identified differentially methylated regions (DMRs) directly from ONT methylation data and built classifiers using a nested cross-validation framework.

**Results:** We identified 143 DMRs for *SETD1B*-related neurodevelopmental disorder (*SETD1B*-NDD), 20 for Kabuki syndrome, and 23 for Kleefstra syndrome; 15 to 26% of the DMRs across the three signatures contained no array probe(s). Cross-validated classifiers ranked every case above every control for all three conditions, and the final classifiers called all but one held-out case correctly, with the remaining case, which carries a *KMT2D* missense variant, as indeterminate. At two outside sites, the classifier for *SETD1B*-NDD correctly categorized seven out of seven cases. Principal component analysis reveals most PacBio and Illumina 5-base genomes cluster with ONT genomes from the same condition. One PacBio control fell between the other controls and the Kabuki cases, whereas the ONT and 5-base genomes for the same individual grouped with controls, suggesting platform-specific methylation differences that the current training cohort is too small to fully account for.

**Conclusions:** Episignatures can be derived directly from sequencing data and applied across platforms. EpiSignaLR is an open workflow that combines the three ONT-derived classifiers with 34 published array-derived signatures. Developing robust sequencing-derived episignatures will require larger cohorts per condition and openly shared, platform-matched methylation data.

## INTRODUCTION

A growing subset of Mendelian conditions, predominantly those caused by disruption of chromatin and methylation machinery, produce a reproducible, disorder-specific pattern of genome-wide DNA methylation change commonly referred to as “episignatures”. These episignatures have been described for more than 100 conditions and have proven diagnostically consequential in two settings: reclassifying variants of uncertain significance (VUS) in an episignature-associated gene, and identifying a potential diagnosis for individuals in whom no causal variant has been identified after typical genetic testing, including exome and genome sequencing.^1–3^

Today, clinical episignature testing is performed almost exclusively on bisulfite-based methylation microarrays, commercialized as the EpiSign assay. Clinically, this testing is typically ordered as a separate assay, most often after non-diagnostic testing or after identification of an uncertain sequencing result, increasing cost and time to diagnosis. For research use, EpigenCentral offers an open alternative: a free web portal that allows researchers, clinicians, and diagnostic laboratories to classify methylation profiles against published episignatures, assess the pathogenicity of variants, and analyze differential methylation without specialized computational infrastructure.^4^ It has also become a route for sharing new signatures and the data behind them. For example, Wang *et al*. used EpigenCentral to validate a refined Kabuki syndrome signature, which included analysis of methylation values extracted from PacBio genomes.^5^ Resources like this make episignature analysis available outside the laboratories that develop signatures, and they are the model we aim to follow for sequencing data.

Array-based testing is inherently limited by the number of CpG sites that can be evaluated. Using arrays, episignatures can only be built from the ∼930,000 CpGs on the array, roughly 3% of CpGs in the genome, so repetitive regions, structurally variable regions, and any region not represented on the array are excluded. In addition, array-based approaches do not allow for phasing of variants or methylation. Thus, when an array-derived signature is applied to genome sequencing data, ∼97% of CpGs are unused. If disorder-specific methylation changes occur outside array sites, signatures discovered directly from genome-wide sequencing data could capture this additional signal, likely improving classification for some conditions and making it possible to build signatures for conditions that lack one on the array.

Long-read sequencing (LRS) is an emerging approach that captures methylation information natively during sequencing. A single ONT or PacBio genome yields single-nucleotide variants, structural variants, tandem repeat expansions, and base modifications at essentially all ∼28 million autosomal CpGs, and allows phasing of these variants. Separately, Illumina 5-base sequencing reports CpG methylation alongside genotypes from a single short-read library.^6^ Previous work has shown that ONT-derived methylation is concordant with microarray-based episignatures and that support vector machine (SVM) classifiers trained on array-defined loci can be used to detect episignatures in samples sequenced on the ONT platform.^7^ Separately, two recent studies applied array-derived classifiers to ONT and PacBio genomes from individuals with Kabuki syndrome and other conditions known to have an underlying methylation signature and also found results concordant with array testing.^5,8^ Importantly, both observed a systematic offset between array and sequencing methylation values, which suggested that sequencing data be included to improve classifier training or replace array-derived data completely.

Here we describe EpiSignaLR, an open resource for episignature classification from methylation-aware genome sequencing, and apply it to three conditions. We perform genome-wide differentially methylated region (DMR) discovery directly from ONT-derived methylation data for three chromatinopathies with established array-derived episignatures: *SETD1B*-related neurodevelopmental disorder (*SETD1B*-NDD), Kabuki syndrome (*KMT2D/KDM6A*), and Kleefstra syndrome (*EHMT1*). Using these data we developed condition-specific classifiers that can be used to evaluate genome-wide sequencing data. We then applied the same classifiers to PacBio and Illumina 5-base data, and compared the results with those of 34 published array-derived classifiers (hereafter referred to as NSBEpi) applied to the same genomes.^7^ EpiSignaLR packages both sets of classifiers in a single workflow that takes an ONT or PacBio alignment or an Illumina 5-base methylation gVCF and returns a classification without additional laboratory work. All methylation data and methods used to develop EpiSignaLR are made publicly available so that other researchers may build on these results, as limited access to underlying methylation data and classification methods have constrained our understanding of methylation differences in rare genetic conditions.

## METHODS

### Participants and ethics approval

Except where noted, individuals were recruited under protocol 20161 (University of Washington, Genomic Discovery Initiative), and all participants or their legal guardians provided written consent. Dried blood spot samples from five individuals with Kabuki syndrome were obtained from the Combined Brain biorepository (combinedbrain.org); analysis of these de-identified samples was determined exempt by the University of Washington IRB (exemption #35386). Five held-out individuals with *SETD1B*-NDD were contributed by the University of British Columbia (UBC) and consented under UBC Research Ethics Board protocol H08-00784; DNA was extracted from whole blood collected in EDTA tubes. All but one of the remaining *SETD1B*-NDD cases were residual, blood-derived DNA that had previously been collected with informed consent for routine diagnostic testing and isolated following routine diagnostic laboratory procedures at Erasmus MC. The use of genome-wide technologies for diagnostic purposes was approved by the Erasmus MC institutional review board (MEC-2012-387). Samples analyzed at the UMC Utrecht were residual DNA specimens remaining after completion of diagnostic testing, which were used for test development and validation. These specimens were analyzed without identifying information, no additional material was collected from patients for this study, and in accordance with institutional policies governing the use of residual clinical specimens, institutional review board approval was not required.

### Sample collection and DNA extraction

Blood was collected either by standard venipuncture into EDTA tubes or as capillary blood with Tasso+ devices (Tasso, Inc.). For both sample types, DNA was extracted within 5 days of collection from 500 µL of blood with the Monarch HMW DNA Extraction Kit for Cells & Blood (New England Biolabs, T3050L) according to the manufacturer’s protocol, except that the vortexing steps during erythrocyte lysis were omitted to limit DNA shearing. At University Medical Center Utrecht (UMC Utrecht), DNA was extracted from peripheral blood collected in EDTA tubes with the chemagic system (Revvity).

Dried blood spots were extracted with the QIAamp DNA Mini Kit (Qiagen, 51306) using a modified protocol. Three 6–7 mm punches per sample were soaked overnight at 37°C in 120 µL Tris-EDTA (TE) buffer and then digested with 280 µL Buffer ATL (Qiagen, 1014785) and 20 µL Proteinase K (New England Biolabs, P8107S) at 56°C for 1 h with shaking at 900 rpm. Buffer AL (300 µL) was added, samples were incubated at 70°C for 10 min, and 150 µL of 100% ethanol was added before the lysate was loaded onto a QIAamp spin column. Columns were washed with 500 µL Buffer AW1, 700 µL Buffer AW2, and 700 µL of 100% ethanol, dried by centrifugation at 20,000 × g for 3 min, and left open at room temperature for 10 min. DNA was eluted in 50 µL Buffer AE prewarmed to 70°C, and the eluate was passed over the column a second time to increase yield.

DNA was quantified using a Qubit 4 Fluorometer (Invitrogen, Q33226) and dsDNA high-sensitivity assay (Invitrogen, Q33231). DNA quality was assessed using a Nanodrop One (Thermo Scientific, 13400518) and fragment size was evaluated using a FEMTO Pulse System (Agilent, M5330AA).

### ONT library preparation, sequencing, and data processing

Libraries sequenced at the University of Washington were prepared with Ligation Sequencing Kit XL V14 (SQK-LSK114-XL; Oxford Nanopore Technologies) and NEBNext reagents (New England Biolabs). Genomic DNA (3.5–5 µg in 80 µL TE) was repaired and end-prepped with 11.7 µL NEBNext FFPE DNA Repair Buffer v2, 3.3 µL NEBNext FFPE DNA Repair Mix, and 5 µL NEBNext Ultra II End-prep Enzyme Mix at 20°C for 30 min and 65°C for 5 min, with the buffer volume scaled for inputs above 80 µL. Thermolabile Proteinase K (4 µL, New England Biolabs) was then added with 26 µL Q5 Reaction Buffer and incubated at 37°C for 10 min and 55°C for 10 min. DNA was purified with a 1x volume of AMPure XP beads (Beckman Coulter), washed twice with 80% ethanol, and eluted in 61 µL TE. Adapters were ligated to 60 µL of end-prepped DNA with 25 µL Ligation Buffer, 10 µL Salt-T4 DNA Ligase (New England Biolabs, M0467), and 5 µL Ligation Adapter for 1 h at room temperature. Libraries were purified with 40 µL AMPure XP beads, washed twice with 250 µL Long Fragment Buffer, and eluted in 97 µL Elution Buffer. DNA was quantified after each cleanup with a Qubit 4 Fluorometer (Invitrogen). Libraries were sequenced on a PromethION 24 using R10.4.1 PromethION or PromethION Plus flow cells to approximately 30x depth of coverage **(Table 1)**.

**Table 1.** Individuals and genomes included in this study. Individuals are grouped by diagnosis with those contributed by UBC and UMC Utrecht listed separately. Gene and variant type refer to the causal variant; controls listed as Other carry a molecular diagnosis in a gene unrelated to the three conditions. Sample indicates venous blood, capillary blood collected with a Tasso device, or a dried blood spot (DBS). ONT set indicates whether an ONT genome was used to train the sequencing-derived classifiers (Training), held out from training (Testing), or sequenced and analyzed externally (External). Coverage is mean genome-wide depth, and N50 is the read N50. –, not sequenced on that platform; NR, not reported.

| Individual | Sex | Age (y) | Gene | Variant type | Sample | ONT set | ONT cov. (x) | ONT N50 (kb) | PacBio cov. (x) | PacBio N50 (kb) | 5-base cov. (x) |
| --- | --- | --- | --- | --- | --- | --- | --- | --- | --- | --- | --- |
| **Controls (n = 23)** |  |  |  |  |  |  |  |  |  |  |  |
| Control-01 | XY | 11–20 | – | – | Capillary | Testing | 32.2 | 30.4 | – | – | 38.1 |
| Control-02 | XX | 41–50 | – | – | Capillary | – | – | – | – | – | 38.2 |
| Control-03 | XX | 0–10 | – | – | Capillary | Training | 40.6 | 11.4 | – | – | 38.7 |
| Control-04 | XY | 11–20 | – | – | Capillary | Training | 44.3 | 10.0 | – | – | 54.3 |
| Control-05 | XY | 11–20 | – | – | Capillary | Testing | 36.5 | 9.8 | – | – | 47.3 |
| Control-06 | XY | 0–10 | – | – | Capillary | – | – | – | – | – | 54.5 |
| Control-07 | XY | 11–20 | – | – | Capillary | – | – | – | – | – | 27.4 |
| Control-08 | XX | 0–10 | – | – | Capillary | Training | 30.7 | 12.9 | 44.8 | 18.3 | 91.6 |
| Control-09 | XX | 41–50 | – | – | Capillary | Training | 30.7 | 13.4 | – | – | 35.1 |
| Control-10 | XY | 41–50 | – | – | Capillary | Testing | 28.8 | 11.9 | 44.8 | 17.7 | 17.9 |
| Control-11 | XX | 0–10 | – | – | Capillary | Testing | 36.4 | 16.9 | 47.5 | 18.3 | 20.1 |
| Control-12 | XY | 0–10 | Other | – | Venous | Training | 32.5 | 33.8 | – | – | – |
| Control-13 | XY | 11–20 | – | – | Venous | Training | 37.3 | 37.3 | – | – | – |
| Control-14 | XY | 0–10 | Other | – | Venous | Training | 31.2 | 21.9 | – | – | – |
| Control-15 | XY | 11–20 | Other | – | Venous | Training | 30.8 | 27.3 | – | – | – |
| Control-16 | XX | 0–10 | Other | – | Capillary | Training | 32 | 33.3 | – | – | – |
| Control-17 | XX | 11–20 | Other | – | Capillary | Training | 32.5 | 29.3 | – | – | – |
| Control-18 | XY | 0–10 | – | – | Capillary | Training | 34.3 | 9.2 | – | – | – |
| Control-19 | XY | 21–30 | – | – | Capillary | Training | 39 | 29.0 | – | – | – |
| Control-20 | XY | 0–10 | – | – | Venous | Training | 31.9 | 17.6 | – | – | – |
| Control-21 | XX | 0–10 | – | – | Capillary | Training | 26.5 | 31.8 | – | – | – |
| Control-22 | XX | 41–50 | – | – | Capillary | Training | 24.3 | 27.5 | – | – | – |
| Control-23 | XY | 11–20 | – | – | Capillary | Training | 25.7 | 29.4 | – | – | – |
| **SETD1B-NDD (n = 11)** |  |  |  |  |  |  |  |  |  |  |  |
| SETD1B-01 | XY | 0–10 | SETD1B | missense | Venous | Training | 24.8 | 7.1 | 35.1 | 15.3 | 144.5 |
| SETD1B-02 | XY | 0–10 | SETD1B | stop | Venous | Training | 21.4 | 11.4 | 46.9 | 16.4 | 79.5 |
| SETD1B-03 | XX | 0–10 | SETD1B | deletion | Venous | Training | 23 | 11.0 | – | – | 137.8 |
| SETD1B-04 | XX | 0–10 | SETD1B | frameshift | Venous | Training | 22.1 | 8.1 | – | – | – |
| SETD1B-05 | XX | 0–10 | SETD1B | missense | Venous | Training | 22.2 | 9.8 | – | – | – |
| SETD1B-06 | XY | 21–30 | SETD1B | in-frame indel | Venous | Training | 25 | 8.0 | 41.2 | 17.2 | 133.4 |
| SETD1B-07 | XY | 0–10 | SETD1B | missense | Venous | Training | 18.2 | 9.2 | – | – | 43.5 |
| SETD1B-08 | XX | 0–10 | SETD1B | deletion | Venous | Training | 18.4 | 3.9 | – | – | 42.1 |
| SETD1B-09 | XY | 31–40 | SETD1B | stop | Venous | Training | 20.3 | 9.3 | – | – | 142 |
| SETD1B-10 | XX | 0–10 | SETD1B | missense | Venous | Training | 19.6 | 7.0 | – | – | – |
| SETD1B-11 | XY | 0–10 | SETD1B | missense | Venous | Testing | 19.7 | 9.1 | – | – | – |
| **Kabuki syndrome (n = 12)** |  |  |  |  |  |  |  |  |  |  |  |
| Kabuki-01 | XX | 0–10 | KMT2D | translocation | Venous | Testing | 33 | 28.6 | 46.4 | 18.3 | 39.6 |
| Kabuki-02 | XX | 31–40 | KMT2D | missense | Capillary | Training | 35.5 | 10.6 | – | – | 36.2 |
| Kabuki-03 | XY | 11–20 | KMT2D | frameshift | Capillary | Training | 31.6 | 14.0 | 46.4 | 17.5 | 146.1 |
| Kabuki-04 | XX | 0–10 | KMT2D | nonsense | Capillary | Training | 33.7 | 9.2 | 37.6 | 15.6 | 94.5 |
| Kabuki-05 | XX | 11–20 | *KMT2D* | missense | Capillary | Training | 34.9 | 13.3 | 48.3 | 18.7 | 41.6 |
| Kabuki-06 | XY | 11–20 | *KMT2D* | frameshift | Capillary | Training | 32.3 | 11.7 | – | – | 35.4 |
| Kabuki-07 | XX | 11–20 | *KDM6A* | nonsense | DBS | Training | 32.3 | 3.2 | – | – | – |
| Kabuki-08 | XX | 0–10 | *KDM6A* | splice | DBS | Training | 27.5 | 6.6 | – | – | – |
| Kabuki-09 | XY | 31–40 | *KMT2D* | missense | DBS | – | – | – | – | – | 130.1 |
| Kabuki-10 | XY | 0–10 | *KMT2D* | nonsense | DBS | Training | 19.7 | 3.3 | – | – | – |
| Kabuki-11 | XX | 0–10 | *KMT2D* | frameshift | DBS | Training | 16.3 | 2.9 | – | – | – |
| Kabuki-12 | XY | 11–20 | *KMT2D* | nonsense | Venous | – | – | – | 42.8 | 18.2 | 71.2 |
| **Kleefstra syndrome (n = 14)** |  |  |  |  |  |  |  |  |  |  |  |
| Kleefstra-01 | XX | 21–30 | *EHMT1* | translocation | Capillary | Testing | 30.6 | 15.5 | – | – | 157.8 |
| Kleefstra-02 | XX | 11–20 | *EHMT1* | deletion | Capillary | Training | 38.5 | 18.2 | 46.5 | 19.9 | 148.4 |
| Kleefstra-03 | XX | 0–10 | *EHMT1* | deletion | Capillary | Training | 40.8 | 13.8 | 43.9 | 17.7 | 165.7 |
| Kleefstra-04 | XY | 0–10 | *EHMT1* | deletion | Capillary | Training | 41.4 | 13.1 | – | – | – |
| Kleefstra-05 | XY | 11–20 | *EHMT1* | deletion | Capillary | Training | 40.6 | 17.9 | 45.8 | 18.1 | 147.8 |
| Kleefstra-06 | XY | 0–10 | *EHMT1* | deletion | Capillary | Training | 45 | 13.0 | – | – | – |
| Kleefstra-07 | XY | 11–20 | *EHMT1* | splice | Capillary | Testing | 36.7 | 15.8 | – | – | 148.9 |
| Kleefstra-08 | XY | 21–30 | *EHMT1* | nonsense | Capillary | Training | 31.9 | 14.6 | 46.3 | 18.1 | 142.9 |
| Kleefstra-09 | XX | 11–20 | *EHMT1* | deletion | Capillary | Training | 37 | 17.9 | – | – | 168.2 |
| Kleefstra-10 | XY | 41–50 | *EHMT1* | missense | Capillary | Training | 37 | 14.5 | – | – | – |
| Kleefstra-11 | XX | 0–10 | *EHMT1* | deletion | Capillary | Training | 37.3 | 14.5 | – | – | – |
| Kleefstra-12 | XX | 31–40 | *EHMT1* | deletion | Capillary | Training | 36.2 | 14.6 | – | – | 187.6 |
| Kleefstra-13 | XX | 11–20 | *EHMT1* | nonsense | Capillary | Testing | 40.2 | 21.0 | – | – | 133.6 |
| Kleefstra-14 | XX | 31–40 | *EHMT1* | nonsense | Capillary | Testing | 41.7 | 14.0 | – | – | 163.2 |
| **UBC (n = 5)** |  |  |  |  |  |  |  |  |  |  |  |
| SETD1B-12 | NR | NR | *SETD1B* | NR | Venous | External | 39 | 11.2 | – | – | – |
| SETD1B-13 | NR | NR | *SETD1B* | NR | Venous | External | 46 | 13.7 | – | – | – |
| SETD1B-14 | NR | NR | *SETD1B* | NR | Venous | External | 29 | 13.2 | – | – | – |
| SETD1B-15 | NR | NR | *SETD1B* | NR | Venous | External | 31 | 13.8 | – | – | – |
| SETD1B-16 | NR | NR | *SETD1B* | NR | Venous | External | 34 | 13.5 | – | – | – |
| **UMC Utrecht (n = 9)** |  |  |  |  |  |  |  |  |  |  |  |
| UMCU-053 (1) | NR | NR | *EHMT1* | NR | Venous | External | 23.1 | 13.1 | – | – | – |
| UMCU-053 (2) | NR | NR | *EHMT1* | NR | Venous | External | 27.0 | 13.3 | – | – | – |
| UMCU-181 | NR | NR | *EHMT1* | NR | Venous | External | 48.2 | 15.3 | – | – | – |
| UMCU-216 | NR | NR | *EHMT1* | NR | Venous | External | 61.6 | 6.7 | – | – | – |
| UMCU-922 | NR | NR | *KMT2D* | NR | Venous | External | 78.6 | 5.4 | – | – | – |
| UMCU-725 | NR | NR | *KMT2D* | NR | Venous | External | 68.4 | 19.7 | – | – | – |
| UMCU-140 | NR | NR | *KDM6A* | NR | Venous | External | 36.8 | 5.4 | – | – | – |
| UMCU-492 | NR | NR | *KDM6A* | NR | Venous | External | 12.2 | 4.6 | – | – | – |
| UMCU-025 | NR | NR | *SETD1B* | NR | Venous | External | 58.7 | 13.9 | – | – | – |
| UMCU-244 | NR | NR | *SETD1B* | NR | Venous | External | 74.5 | 6.1 | – | – | – |

Raw signal data were basecalled with Dorado v1.1.1 and v1.3.0 (ONT) using the v5.2 SUP model with 5mCG and 5hmCG modification calling (dna_r10.4.1_e8.2_400bps_sup@v5.2.0_5mCG_5hmCG@v2). Reads were aligned to the GRCh38 reference genome with minimap2 (v2.28-r1209).^9^ Single-nucleotide variants and small insertions and deletions were called with Clair3 (v1.0.8)^10^ using Rerio model r1041_e82_400bps_sup_v520 (https://github.com/nanoporetech/rerio), and structural variants were called with Sniffles2 (v2.8).^11^ Small variants were phased and structural variants co-phased with LongPhase (v1.7.3),^12^ and reads were haplotagged for the phased methylation pileups described below.

Libraries sequenced at UMC Utrecht were prepared, sequenced, and processed with a separate protocol. DNA was fragmented by FastPrep homogenization (MP Biomedicals), and libraries were prepared with the Ligation Sequencing Kit XL V14 (SQK-LSK114-XL) using the standard whole-genome protocol on a Callisto sample preparation system (Volta Labs). Each sample was sequenced on a PromethION 24 using two R10.4.1 flow cells (FLO-PRO114M) for 72 hours without washing, targeting more than 30x combined coverage; three genomes from two individuals fell short of this target **(Table 1)**. Basecalling with the dna_r10.4.1_e8.2_400bps_HAC@v5.2.0 model, including 5mC and 5hmC calling in CpG contexts, and alignment to GRCh38 were performed during sequencing in MinKNOW (v26.01.15). Data from the two flow cells were combined and processed with the EPI2ME wf-human-variation workflow (v2.7.2; https://github.com/epi2me-labs/wf-human-variation), and the resulting alignments were classified with EpiSignaLR using default settings, applying the array-derived classifiers and the sequencing-derived *SETD1B*-NDD classifier. These genomes were classified with an earlier release of the *SETD1B*-NDD model, which used 30 DMRs rather than the 23 in the final model described below.

### PacBio HiFi library preparation and sequencing

Fifteen individuals **(Table 1)** were sequenced on a PacBio Revio system with SPRQ-Nx chemistry. Libraries were barcoded and pooled in groups of three, and each pool was sequenced across three SMRT Cell acquisitions, for an average of one SMRT Cell per sample. HiFi reads were generated on-instrument with Revio software v13.5.0, which infers CpG 5mC from polymerase kinetics with jasmine v26.1.3 and stores per-read modification probabilities in the MM and ML tags of each read. Reads from the three acquisitions of each sample were aligned to GRCh38 with pbmm2 and merged. Per-CpG methylation was then summarized with the aligned_bam_to_cpg_scores tool from pb-CpG-tools (v3.0.0)^13^ in count mode (--pileup-mode count --modsites-mode reference), which classifies each read at each reference CpG as methylated or unmethylated, combines the two strands, and reports the fraction of methylated reads and the read depth.

### Illumina 5-base library preparation and sequencing

Illumina 5-base libraries were prepared according to the Illumina 5-Base DNA Prep product documentation (document 200073497 v01).^6^ Genomic DNA was fragmented by sonication to an average size of approximately 450 bp, end-repaired, A-tailed, and ligated to UMI adapters. Methylated cytosines were then converted to thymine in a single enzymatic step, leaving unmethylated cytosines unchanged, and libraries were amplified with unique dual indexes. Libraries were sequenced on a NovaSeq X Plus using 25B flow cells with 2×151-bp paired-end reads. Reads were analyzed with the DRAGEN v4.5 Germline 5-base pipeline^14^ using GRCh38. The pipeline aligns reads with an aligner adjusted for converted bases and identifies methylation from C>T mismatches on the forward strand and G>A mismatches on the reverse strand; because a methylation event appears on only one strand whereas a genetic variant is supported by both, methylation and sequence variants are called from the same reads. CpG methylation was reported in the gVCF as the fraction of modified calls and the informative read depth, aggregated across the two strands of each CpG.

### Differentially methylated region discovery

For ONT data, methylation pileups were generated using modkit v0.6.1^15^ with the parameters --modified-bases 5mC 5hmC --cpg --combine-strands --phased --combine-mods --filter-threshold C:0.7 --mod-threshold m:0.7 --mod-threshold h:0.7 to combine the 5mC and 5hmC at canonical CpG sites in the GRCh38 reference genome. The haplotype agnostic “combined” pileup was then filtered for CpGs not overlapping centromeric regions, segmental duplications, simple repeats over 10,000 bp, gaps, and common CNVs within the canonical autosomes. Haplotype-resolved pileups produced with --phased were not used for classification.

dmrseq was used to determine differentially methylated regions between cases and controls. ^16,17^ For each condition, 10 cases (9 for Kabuki syndrome) and 10 controls, matched for age as closely as available samples allowed, were selected for the discovery/training cohort and filtered for CpGs in high confidence regions with at least 5 valid reads. The number of valid reads and the number of modified reads at each position for each sample was formatted into a BSseq object for dmrseq. The age and sex of each sample were used as covariates to account for other contributors of methylation variation. Significant DMRs were filtered for an adjusted p-value (q-value) of 0.05 or less **(Supplemental Tables 1–3)**. Controls for each condition were drawn from a pool of 16 individuals without a variant in *SETD1B*, *KMT2D*, *KDM6A*, or *EHMT1* **(Table 1)**. Eleven had no reported genetic diagnosis, and five had a molecular diagnosis in a gene unrelated to these conditions, meaning the control pool includes individuals with other genetic conditions as well as unaffected individuals.

### Methylation classifier feature selection and training

The average coverage-weighted methylation fraction was calculated for each DMR for each sample in the discovery/training cohort for feature selection. Given the small sample size, scaling, feature selection, and hyperparameter tuning were performed within a nested leave-one-out cross-validation (LOOCV) framework to estimate generalization performance and to prevent information from the held-out sample influencing feature selection. DMR discovery itself was performed once on the full discovery cohort before cross-validation. In each outer fold, one sample was withheld and the remaining 19 (18 for Kabuki syndrome) were used for model construction. Features were selected using an elastic-net–penalized logistic regression.^18^ The elastic-net formulation was chosen over a pure LASSO penalty because the L2 component retains groups of correlated DMRs rather than arbitrarily selecting a single representative, yielding a larger and more reproducible feature panel. The regularization strength (*C*) and the L1/L2 mixing parameter (l1_ratio) were tuned by stratified 5-fold cross-validation on the training samples only (grids: *C* ∈ {0.05, 0.1, 0.25, 0.5, 1.0}, l1_ratio ∈ {0.3, 0.5, 0.7, 0.9}); a minimum of 5 and a maximum of 23 features were retained per fold, ranked by absolute coefficient when the number of nonzero coefficients fell outside these bounds. All DMR features were standardized (zero mean, unit variance) before feature selection, using parameters estimated from the training samples alone.

A linear-kernel SVM was then trained on the selected, standardized features, with its regularization parameter (*C* ∈ {0.01, 0.1, 1, 10}) tuned in the same inner cross-validation. Classifier performance was quantified by pooling the held-out predictions across all outer folds and computing the area under the receiver operating characteristic curve (AUC), balanced accuracy, sensitivity, and specificity. Statistical significance was assessed by a label-permutation test with 200 permutations, in which the full nested LOOCV procedure was repeated on randomly shuffled class labels to construct a null distribution of AUC values; the permutation p-value was calculated as (*b* + 1)/(*n* + 1), where b is the number of permuted AUCs greater than or equal to the observed AUC and *n* = 200, so the smallest attainable *p* value is 0.005. The training-set fit of the final model was not used as a performance estimate.

Feature-panel stability was assessed in two ways: the frequency with which each DMR was selected across the outer folds, and the frequency of selection across 500 bootstrap resamples of the full cohort, with DMRs selected in ≥60% of resamples reported as stable.

The final classifier for deployment was refit on all training samples, with hyperparameters selected by 5-fold cross-validation, and its per-DMR linear weights were retained for interpretation. Class probabilities were obtained via Platt scaling;^19^ samples with a probability of 0.5 or higher are classified as positive, and those from 0.4 to 0.6 are flagged as indeterminate. The final models were applied to three groups of samples. The first comprised individuals who were not used at any stage of DMR discovery, feature selection, training, or hyperparameter tuning for that model: six *SETD1B* cases (one from Erasmus MC and five from the University of British Columbia), one Kabuki case, four Kleefstra cases, and four controls sequenced with ONT, controls from the shared pool who were not selected for that model’s training cohort, cases of the other two conditions, and individuals sequenced only on other platforms **(Table 1)**. The second comprised PacBio and Illumina 5-base genomes from individuals in the ONT training cohorts. These genomes test whether a classifier trained on ONT data can be applied to another platform, but not whether it generalizes to new individuals. The third comprised 10 ONT genomes from nine individuals sequenced at UMC Utrecht **(Table 1)**, none of whom contributed to any stage of model development. All analyses were performed in Python 3.14 using scikit-learn v1.8.0.^20^

### EpiSignaLR Workflow

The EpiSignaLR workflow takes an ONT modBAM, a PacBio aligned BAM, or an Illumina 5-base methylation gVCF aligned to GRCh38 and performs three steps: it generates a per-CpG methylation pileup, classifies the sample with 34 array-derived signatures, and classifies the sample with three sequencing-derived signatures **(Figure 1A, Supplemental Table 4)**. ONT pileups are generated with modkit using the parameters described above, PacBio pileups with aligned_bam_to_cpg_scores from pb-CpG-tools (count pileup mode, reference CpG sites), and Illumina 5-base pileups by extracting the per-CpG methylation fraction (M5mC) and informative depth (DPM5mC) from PASS reference blocks of the DRAGEN gVCF. The array-derived classifiers were developed by Geysens *et al*. for NSBEpi and are adapted here from their published code.^7^ The pileup is intersected with the 3,642 CpGs that make up the array-derived episignatures. For each of the 34 conditions, a linear SVM that weights the target condition 10-fold is trained on the reference profiles which give one median methylation profile per condition at each signature’s CpGs, derived from the array study of Aref-Eshghi *et al*. and applied to the sample, with CpGs missing from the sample imputed with the reference mean. The sample is assigned to the highest-scoring condition, or to control if no SVM decision value reaches 0.35; when more than one condition exceeds 0.35, all are reported. For ONT-derived signatures, methylation at each DMR is calculated as a coverage-weighted pooled fraction (modified calls divided by valid coverage, summed across the DMR’s CpGs), using CpGs covered by at least 2 reads for ONT and PacBio or at least 10 informative reads for Illumina 5-base. A DMR with no qualifying CpGs is imputed with the training-cohort mean. A sample is classified as positive at a probability of 0.5 or higher, and scores from 0.4 to 0.6 are flagged as indeterminate. For each model, EpiSignaLR also produces a PCA plot and a clustered heatmap as visual companions placing the sample alongside the training cohort at the selected DMRs.

**Figure 1.**
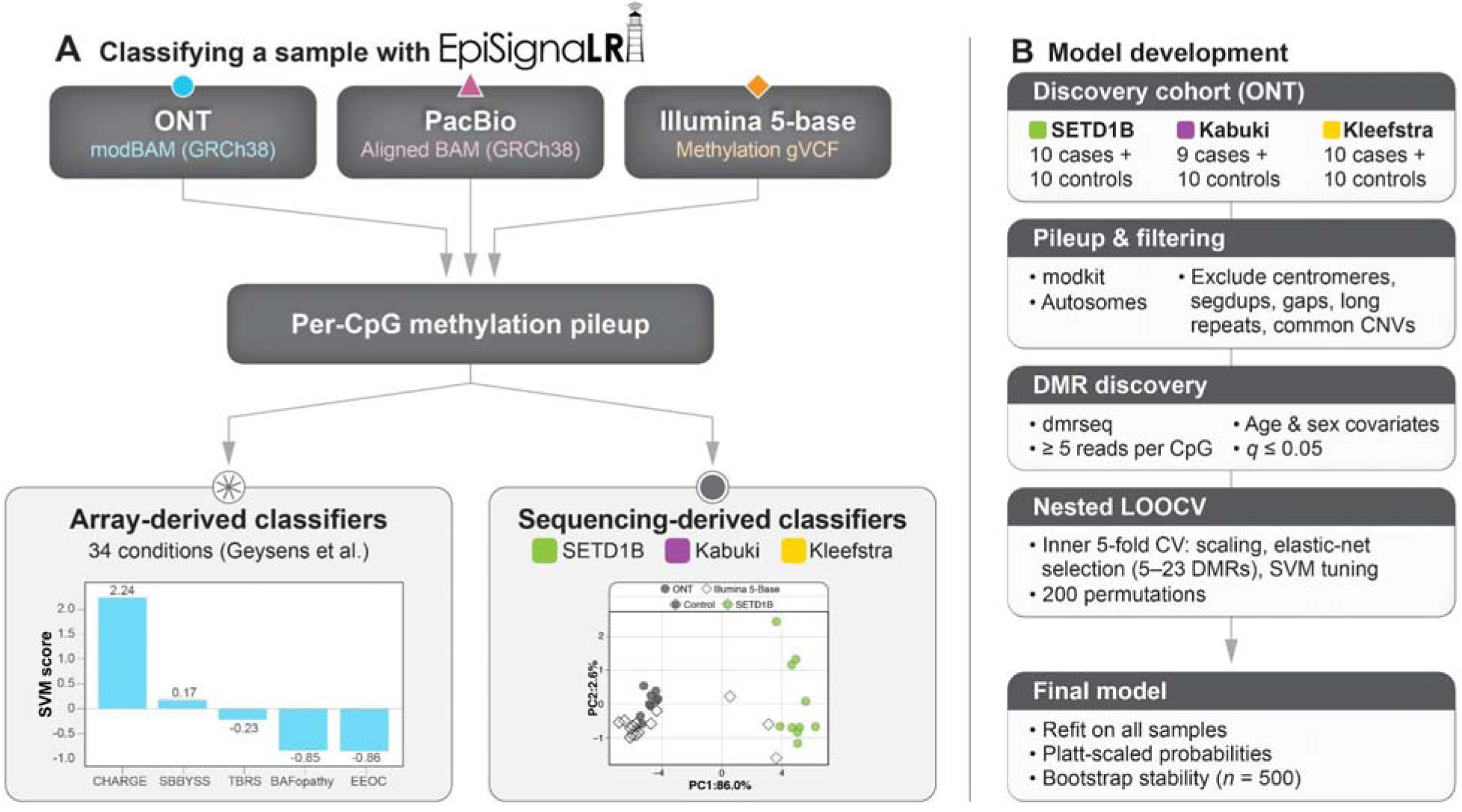
EpiSignaLR workflow. **(A)** A sample is classified from an ONT modBAM, a PacBio aligned BAM, or an Illumina 5-base methylation gVCF. A platform-specific step produces a per-CpG methylation pileup, which is scored by 34 array-derived classifiers adapted from NSBEpi and by three sequencing-derived classifiers. **(B)** Development of each sequencing-derived classifier from an ONT discovery cohort.

## RESULTS

### EpiSignaLR classifies samples from three sequencing platforms

EpiSignaLR is a single command-line workflow that takes an ONT or PacBio aligned BAM, or an Illumina 5-base methylation gVCF aligned to GRCh38 and returns two sets of results **(Figure 1A)**. Each input is first converted to a per-CpG methylation pileup with a platform-specific tool: modkit for ONT,^15^ aligned_bam_to_cpg_scores from pb-CpG-tools for PacBio,^13^ and a parser that extracts the per-CpG methylation fraction and informative depth from DRAGEN methylation gVCFs for Illumina 5-base data. All later steps are shared across platforms. In the first arm, the pileup is intersected with the 3,642 CpGs that make up published array episignatures using bedtools,^21^ and 34 linear SVMs adapted from NSBEpi score the sample against reference methylation profiles for each condition.^7^ In the second arm, methylation is averaged across the DMRs of each sequencing-derived signature and scored with the corresponding elastic-net and SVM classifier, which reports a probability, a call, an indeterminate flag, and a PCA and heatmap placing the sample alongside its training cohort.

EpiSignaLR runs in under 30 minutes on a single compute node. For example, starting from an aligned BAM or methylation gVCF (basecalling, alignment, and variant calling are not included), a complete run with 20 threads on one control individual (Control-10) took 11 minutes for a 29x ONT genome, 24 minutes for a 45x PacBio HiFi genome, and 19 minutes for an Illumina 5-base genome, with peak memory of 27, 15, and 15 GB, respectively.

### Discovery cohorts

We built sequencing-derived signatures from ONT whole-genome sequencing data from individuals with molecularly confirmed *SETD1B*-related neurodevelopmental disorder (*SETD1B*-NDD *n* = 10), Kabuki syndrome (*n* = 9; 7 with *KMT2D* and 2 with *KDM6A* variants), and Kleefstra syndrome (*n* = 10), each compared with 10 controls drawn from a pool of 16 individuals who were either self-reported healthy controls or individuals affected by a known genetic condition without a variant in the target genes **(Figure 1B**, **Table 1)**. Eleven of these controls had no reported genetic diagnosis and five had a molecular diagnosis in an unrelated gene, so each classifier was trained to separate its condition from both unaffected individuals and individuals with other genetic conditions. Median ONT coverage among cases was 21.8x (range 18.2–25.0x) for *SETD1B*-NDD, 32.3x (16.3–35.5x) for Kabuki syndrome, and 37.9x (31.9–45.0x) for Kleefstra syndrome, with median read N50 values of 8.7, 9.2, and 14.6 kb. Controls in the pool had a median ONT coverage of 32.0x (24.3–44.3x) and a median read N50 of 27.4 kb (9.2–37.3 kb). Illumina 5-base libraries were generated for 36 individuals and PacBio HiFi libraries for 15. PacBio libraries had a median coverage of 45.8x (35.1–48.3x) and a median read N50 of 18.1 kb. Illumina 5-base coverage differed between groups: libraries from cases had a median coverage of 137.8x (35.4–187.6x), compared with 38.2x (17.9–91.6x) for controls; because of this difference, we cannot exclude a contribution of coverage to the Illumina 5-base scores. Most of these libraries came from individuals in the ONT training cohorts, including 22 of 36 Illumina 5-base libraries and 11 of 15 PacBio libraries.

### Array-derived classifiers applied to sequencing data

We first asked how well published array-derived classifiers perform on genome sequencing data. We applied the 34 array-derived classifiers in EpiSignaLR to every ONT, PacBio HiFi, and Illumina 5-base genome in the study and called a result positive when its SVM decision value exceeded 0.3 **(Figure 2A–C)**. These classifiers were trained on published median methylation values, one profile per condition, rather than on individual-level array data, so each had a single positive training example. Performance differed by condition. The Kleefstra classifier called all 14 individuals with Kleefstra syndrome on every platform on which they were sequenced (28 of 28 genomes). The Kabuki classifier called all 13 PacBio and Illumina 5-base genomes from individuals with Kabuki syndrome, including Kabuki-09-ILM, but only 8 of 10 ONT genomes. The two it missed came from the two individuals with *KDM6A* variants, and one of them, Kabuki-07, was assigned to Rubinstein-Taybi syndrome (RSTS; 0.49). The *SETD1B*-NDD array-derived classifier performed the worst. It called 9 of 16 ONT genomes (including the five from UBC; see below), 1 of 3 PacBio genomes, and none of the 6 Illumina 5-base genomes from individuals with *SETD1B*-NDD, thus only 10 of 16 individuals were called on any platform, and calls disagreed between platforms for three of the six individuals sequenced on more than one. No control scored above 0.3 on the *SETD1B*-NDD, Kabuki, or Kleefstra classifiers **(Figure 2D–F)**, but several genomes scored above 0.3 on classifiers for other conditions **(Figure 3)**. Nine of the 14 individuals with Kleefstra syndrome also scored above 0.3 for Tatton-Brown-Rahman syndrome (TBRS) on at least one platform, although their Kleefstra score was always higher. One individual with Kabuki syndrome, (Kabuki-01) scored above 0.3 for RSTS on all three platforms and for Say-Barber-Biesecker-Young-Simpson syndrome (SBBYSS) on two; another (Kabuki-02) scored above 0.3 for TBRS on its Illumina 5-base genome. Two controls were also called: Control-08 for TBRS on Illumina 5-base (0.42), and Control-09 for TBRS (0.44) and SBBYSS (0.31) on Illumina 5-base and for SBBYSS on ONT (0.34). No individual with *SETD1B*-NDD scored above 0.3 on another classifier. No *SETD1B*-NDD individuals were included in the validation for NSBEpi, but they reported that all of their Kabuki and Kleefstra individuals classified correctly. Because the array-derived classifiers transferred well for some conditions and poorly for others, we next evaluated the performance of signatures derived directly from sequencing data.

**Figure 2.**
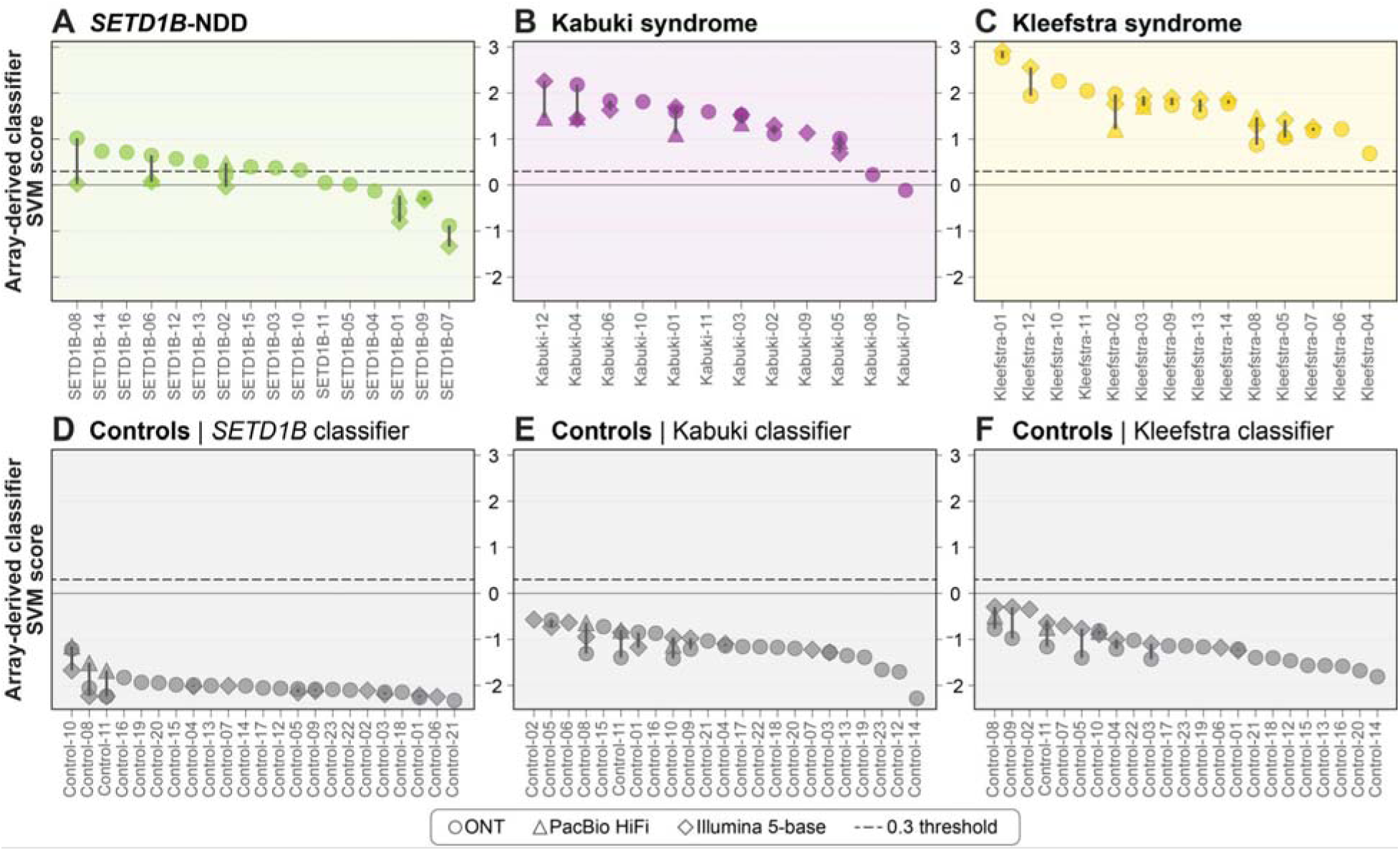
Array-derived episignature scores for cases and controls across sequencing platforms. Each of the 34 array-derived support vector machine (SVM) classifiers adapted from NSBEpi was applied to every genome. Panels show scores from the *SETD1B* **(A, D)**, Kabuki **(B, E)**, and Kleefstra **(C, F)** classifiers for individuals with the corresponding condition **(A–C)** and for controls **(D–F)**. Each column is one individual, ordered by highest score; genomes from the same individual sequenced on more than one platform are plotted in the same column and joined by a vertical line. The dashed line marks the 0.3 threshold above which a condition is called.

**Figure 3.**
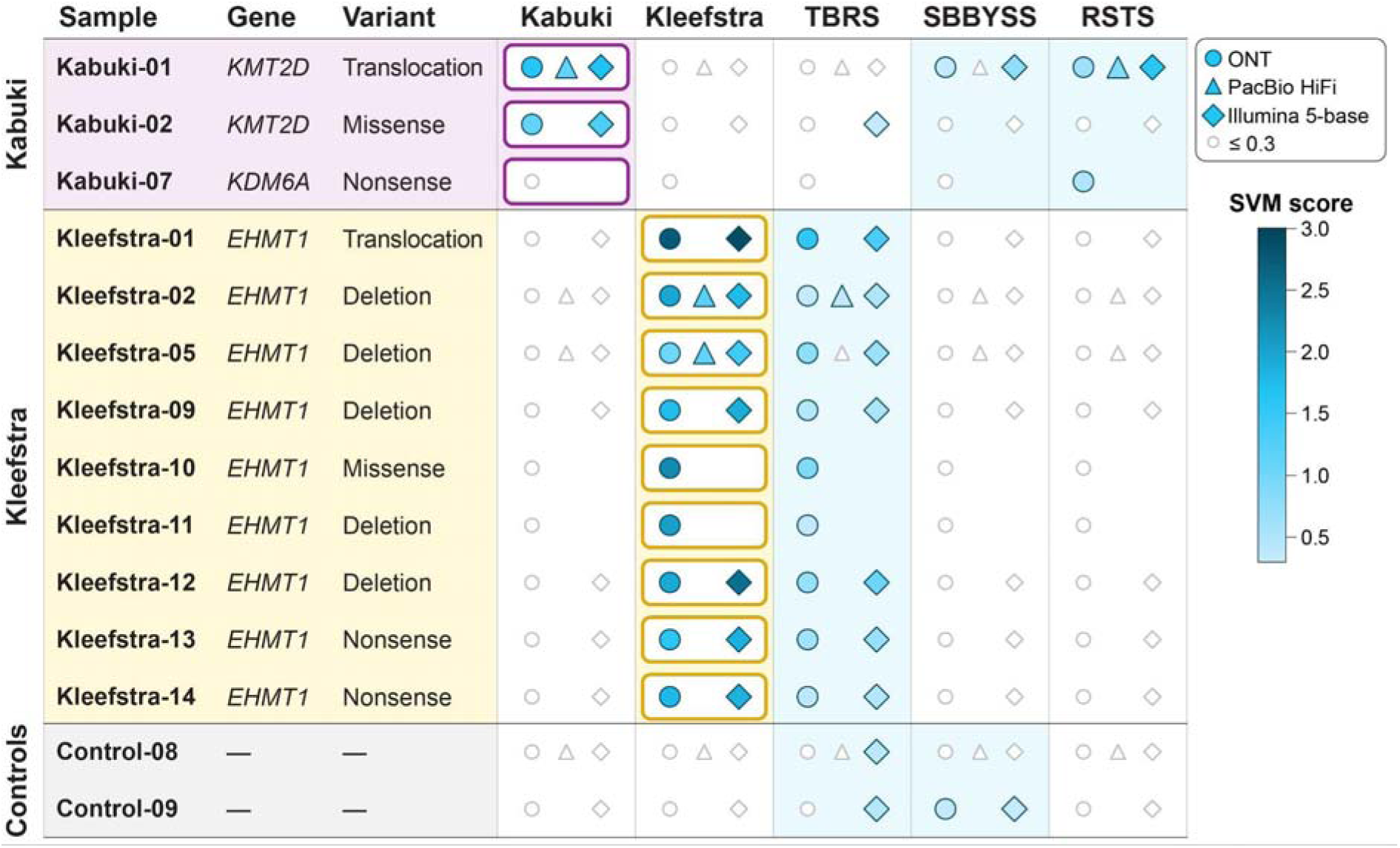
Genomes with array-derived classifier scores above 0.3 for a condition other than their own. Rows are individuals with at least one score above 0.3 from a classifier other than the one for their diagnosis, grouped by diagnosis and labeled with the affected gene and variant type **(Table 1)**. Columns are the array-derived classifiers on which any of these individuals scored above 0.3: Kabuki syndrome, Kleefstra syndrome, Tatton-Brown-Rahman syndrome (TBRS), Say-Barber-Biesecker-Young-Simpson syndrome (SBBYSS), and Rubinstein-Taybi syndrome (RSTS). Filled symbols are colored by SVM score where the score exceeds 0.3, faint outlines mark scores of 0.3 or below, and an empty position indicates that the individual was not sequenced on that platform. No individual with *SETD1B*-NDD scored above 0.3 on another classifier.

### ONT-derived episignatures

Genome-wide DMR discovery with dmrseq identified 143 DMRs for *SETD1B*-NDD, 20 for Kabuki syndrome, and 23 for Kleefstra syndrome at q ≤ 0.05 **(Supplemental Tables 1–3)**. The DMRs had median widths of 606, 568, and 580 bp and contained a median of 18, 30, and 20 CpGs, respectively. The direction of change differed by condition: 120 of 143 *SETD1B*-NDD DMRs (84%) were hypermethylated in cases, whereas 14 of 20 Kabuki DMRs (70%) and 21 of 23 Kleefstra DMRs (91%) were hypomethylated **(Figure 4A)**. Most DMRs overlapped at least one array probe, but 35 (24%), 3 (15%), and 6 (26%) contained none for *SETD1B*-NDD, Kabuki syndrome, and Kleefstra syndrome respectively. Overlap with the CpGs used by the array-derived classifiers for the same conditions was small: 14 of 91 *SETD1B* signature CpGs (in 12 DMRs), 2 of 153 Kabuki signature CpGs, and 5 of 107 Kleefstra signature CpGs fell within the sequencing-derived DMR set. This is similar to previous observations for Kabuki syndrome, where two array signatures from the same group shared only 78 probes yet both classified samples accurately.^5,22^ Together these results suggest that the methylation changes in these conditions are spread across many CpGs, and that any single signature samples a subset of them. Because the two classifier sets share few CpGs, agreement between them for an individual provides largely independent support for the result.

**Figure 4.**
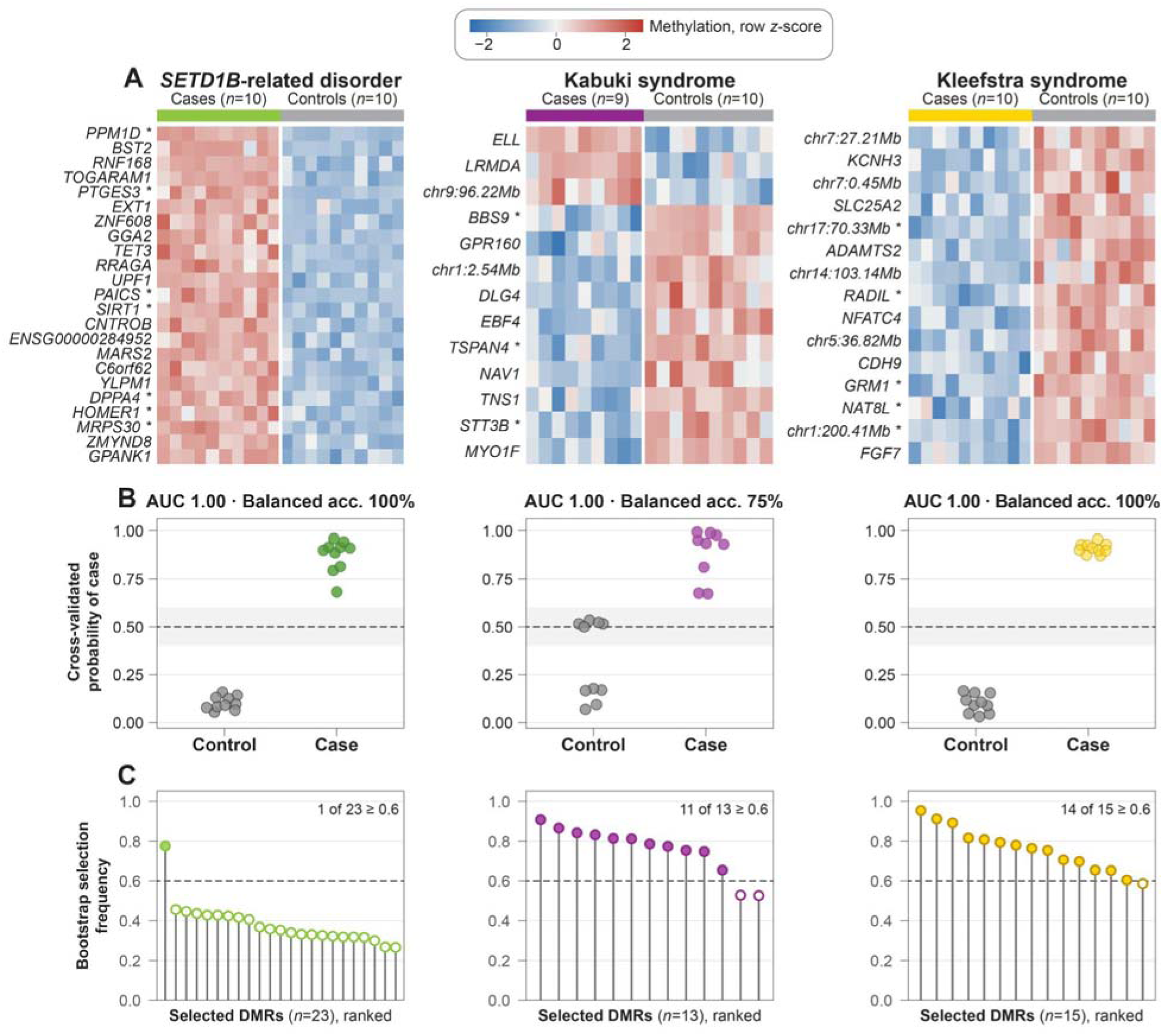
Sequencing-derived episignatures and cross-validated performance. **(A)** Methylation at the DMRs selected by each final model, shown as row z-scores across the training cohort. Rows are ordered by the difference between cases and controls and labeled with the nearest gene, or with genomic position where no gene is annotated. Asterisks mark DMRs that contain no array probe. **(B)** Probability of being a case for each training sample when it was held out in nested leave-one-out cross-validation. The dashed line marks the 0.5 classification threshold and the shaded band the indeterminate zone (0.4–0.6). **(C)** Frequency with which each DMR in the final model was selected across 500 bootstrap resamples; filled points meet the 0.6 stability threshold.

### Classifier performance

Using nested leave-one-out cross-validation, we identified *SETD1B*-NDD, Kabuki, and Kleefstra models that used 23, 13, and 15 DMRs **(Figure 4A)**. Cross-validated predictions ranked every case above every control for all three conditions (AUC 1.00; permutation *p* = 0.005, the smallest value attainable with 200 permutations; **Figure 4B**). For *SETD1B*-NDD and Kleefstra syndrome, the lowest case probabilities were 0.68 and 0.87 and the highest control probabilities were 0.16 in both, giving 100% sensitivity and specificity. For Kabuki syndrome, all nine cases scored 0.67 or higher, but five of 10 controls scored between 0.50 and 0.53, inside the indeterminate zone. At the 0.5 threshold these controls count as false positives, giving a specificity of 50% and a balanced accuracy of 75%. Because the ranking was perfect, this reflects where Platt scaling placed the probability threshold when each cross-validation model was fit on only 18 samples (the 9 cases and 10 controls of the Kabuki training cohort, minus the held-out sample), rather than overlap between groups, and none of these controls received a confident positive call.

Feature stability differed between conditions **(Figure 4C)**. Across 500 bootstrap resamples, 14 of 15 Kleefstra DMRs and 11 of 13 Kabuki DMRs were selected at least 60% of the time, compared with 1 of 23 *SETD1B*-NDD DMRs. Within the leave-one-out folds, 12 of the 23 *SETD1B*-NDD DMRs were selected in every fold, so the instability reflects the large number of correlated *SETD1B*-NDD DMRs that can stand in for one another rather than a weak signal. All three final models selected the strongest regularization in each hyperparameter grid (elastic-net *C* = 0.05, l1_ratio = 0.3, SVM *C* = 0.01).

### Classification across sequencing platforms

Before applying the ONT-trained classifiers to other platforms, we compared per-CpG methylation genome-wide between platforms in individuals sequenced on more than one **(Figure 5)**. Methylation was most concordant between ONT and Illumina 5-base (*r* = 0.955–0.970) and least concordant between ONT and PacBio (*r* = 0.902–0.910). CpGs that were fully methylated on ONT or Illumina 5-base were often reported by PacBio as partially methylated, and no CpG exceeded a PacBio methylation fraction of 0.975.

**Figure 5.**
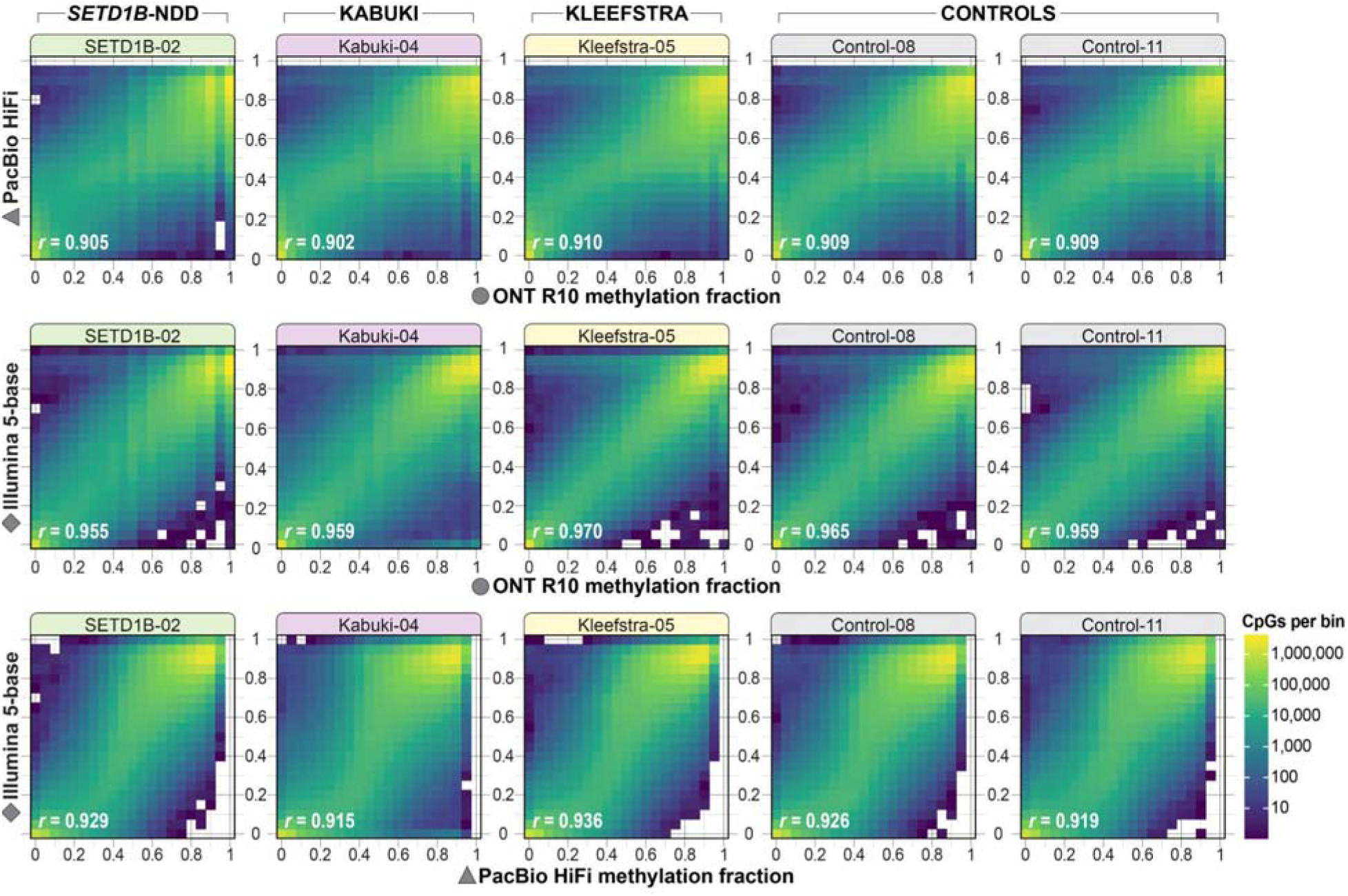
Concordance of CpG methylation across sequencing platforms. Each panel is a two-dimensional histogram comparing per-CpG methylation fractions measured on the same sample from the same individual on two platforms: **(A)** ONT and PacBio HiFi, **(B)** ONT and Illumina 5-base, and **(C)** PacBio HiFi and Illumina 5-base. Columns show one individual with *SETD1B*-NDD (SETD1B-02), one with Kabuki syndrome (Kabuki-04), one with Kleefstra syndrome (Kleefstra-05), and two controls (Control-08 and Control-11). Each tile is a 0.05 × 0.05 bin of methylation fraction, colored by the number of CpGs it contains on a log scale shared across all panels; white tiles contain no CpGs. CpGs were included if they were covered by at least 10 reads on both platforms and not found within telomeres, centromeres, known gaps in GRCh38, common CNVs, or repetitive elements longer than 10,000 bp; r, Pearson correlation coefficient across all included CpGs. Agreement was highest between ONT and Illumina 5-base (*r* = 0.955–0.970) and lowest between ONT and PacBio HiFi (*r* = 0.902–0.910). In comparisons involving PacBio, CpGs that are fully methylated on the other platform are often reported by PacBio as only partially methylated, and no CpG reaches a PacBio methylation fraction above 0.975.

We next evaluated whether classifiers trained on ONT data classify genomes sequenced on other methylation-aware platforms. Because most PacBio and Illumina 5-base genomes came from individuals in the ONT training cohorts, these comparisons mainly test whether the signatures carry over to another platform, while a smaller set of individuals not used in training tests whether they generalize to new individuals. In principal component analysis of average per-DMR methylation, Illumina 5-base and PacBio genomes from cases and controls grouped with the corresponding ONT samples for all three conditions, with one exception described below **(Figure 6A–C)**, and the four groups remained distinct when the DMRs of all three signatures were analyzed together **(Figure 6E)**. Two Kabuki samples that were not used in training, Kabuki-01-ILM and Kabuki-12-ILM, grouped with Kabuki cases **(Figure 6C)**, whereas a third, Kabuki-09-ILM (extracted from a dried blood spot), lay between the Kabuki and control clusters. Kabuki-12 was originally sequenced with ONT R9 chemistry and is therefore represented only by its Illumina 5-base and PacBio data.

**Figure 6.**
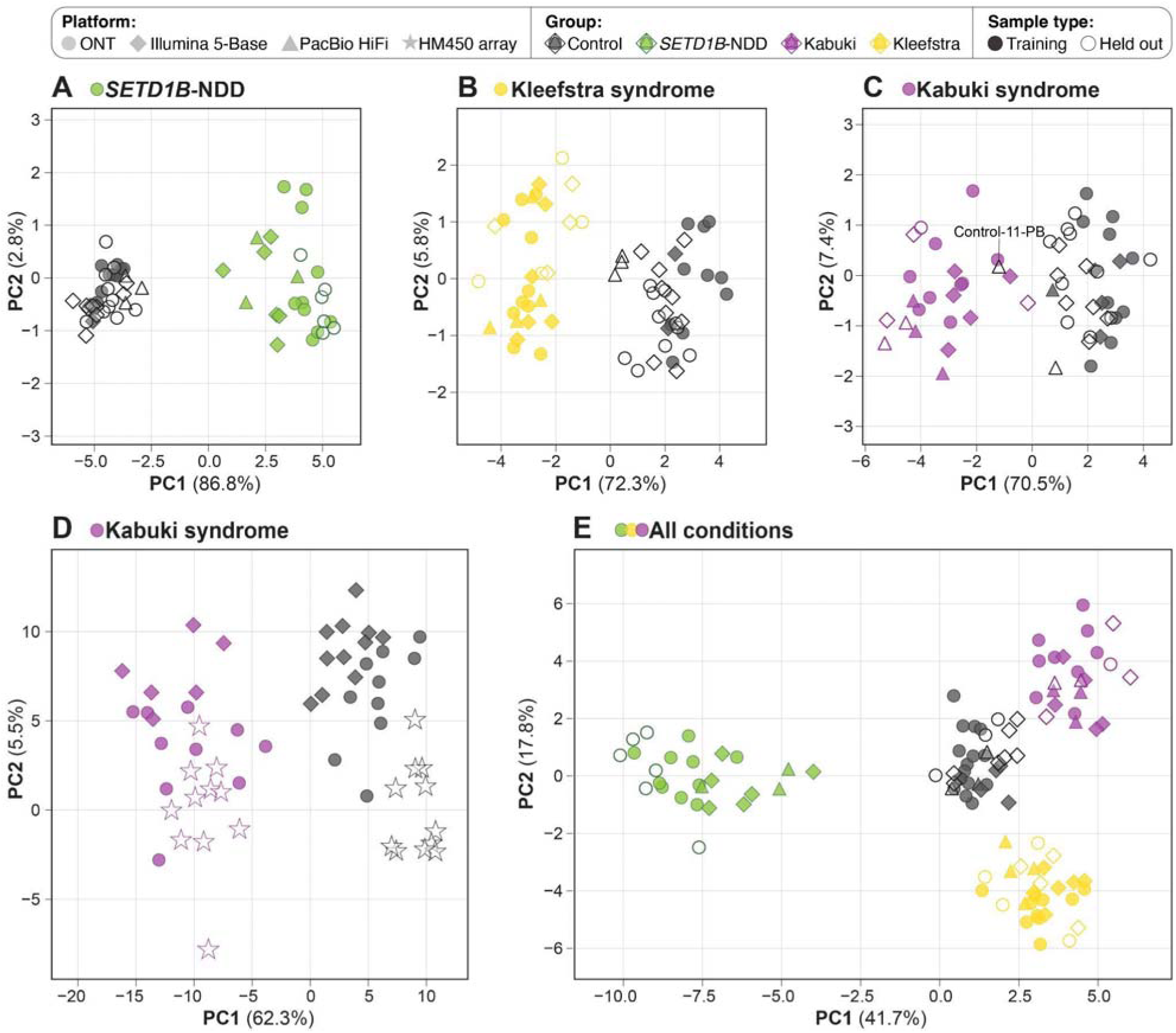
Sequencing-derived episignatures across sequencing platforms. In **A–C** and **E**, filled symbols are individuals in the ONT training cohort, including their PacBio and Illumina 5-base genomes, and open symbols are individuals not used in training. **(A)** Of the six held-out *SETD1B*-NDD ONT genomes, five were externally sequenced. **(B)** Kleefstra syndrome. **(C)** Kabuki syndrome. One PacBio control (Control-11-PB) lies between the control and Kabuki clusters, whereas the ONT and Illumina 5-base genomes from the same individual cluster with controls. **(D)** Kabuki syndrome DMRs in ONT and Illumina 5-base genomes together with public HumanMethylation450 array data from individuals with *KMT2D* variants and controls (stars; **Supplemental Table 5**). In this panel, fill does not indicate training status. PC1 separates cases from controls on all three platforms, and PC2 largely separates samples by platform within each group. **(E)** DMRs from all three signatures analyzed together.

Cross-platform agreement was also visible at individual DMRs. The *PPM1D* DMR, which contains no array probes and was the only *SETD1B*-NDD DMR selected in more than 60% of bootstrap resamples, was hypermethylated in *SETD1B*-NDD cases when compared to controls across all three platforms **(Figure 7)**. In cases, methylation rose abruptly at the boundary of the unmethylated PPM1D promoter region, whereas in controls it rose gradually across the DMR, and this difference was preserved in PacBio genomes despite having higher methylation within the unmethylated region.

**Figure 7.**
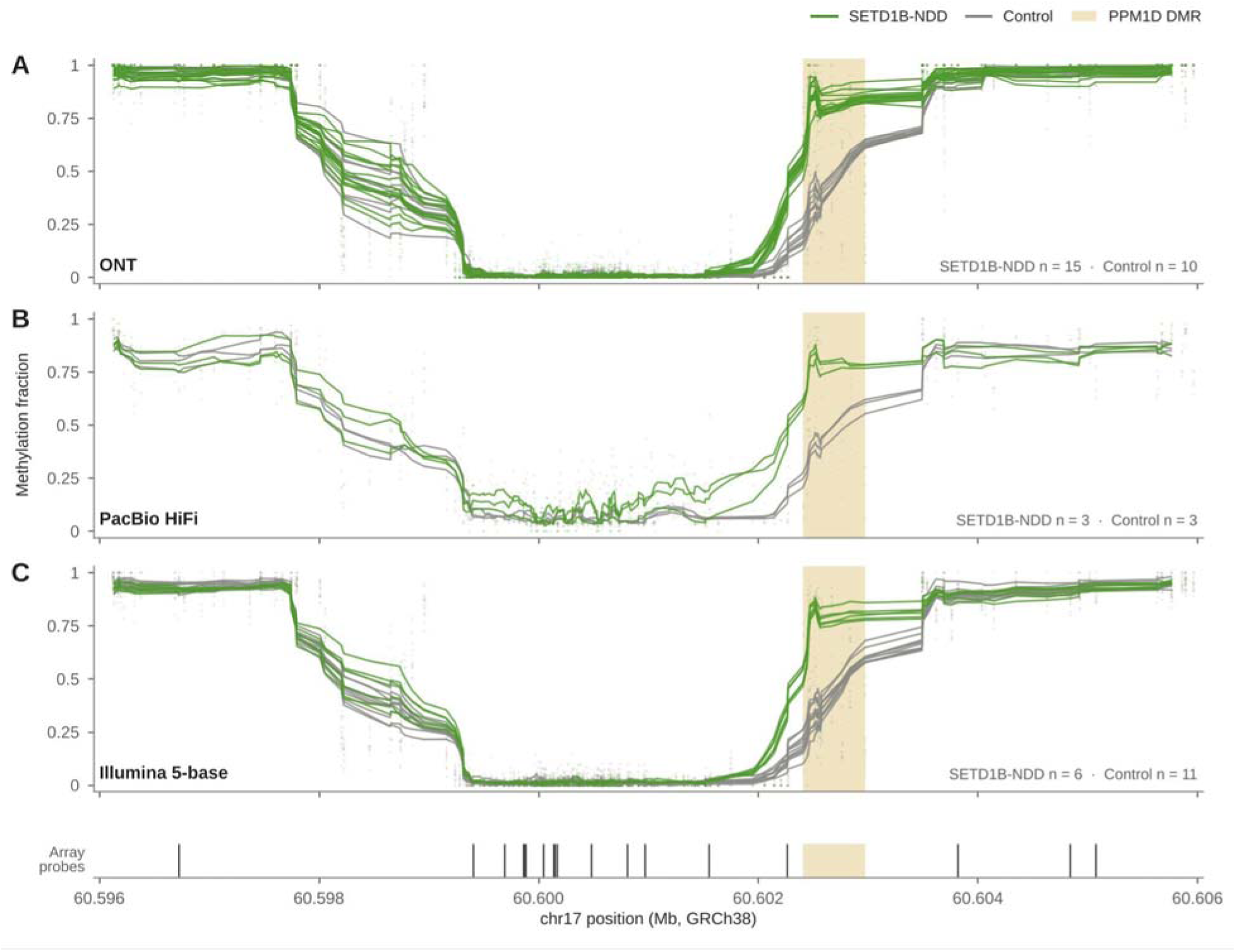
Methylation at the *PPM1D* DMR across sequencing platforms. Methylation fraction across a 10-kb region of chromosome 17 (GRCh38) surrounding the *PPM1D* DMR (chr17:60,602,408–60,602,976; shaded), which lies at the edge of the unmethylated *PPM1D* promoter region. This DMR contains no array probe and is the only *SETD1B*-NDD DMR selected in more than 60% of bootstrap resamples (78%; Figure 4C). Each line is one genome, smoothed across neighboring CpGs, and faint points are methylation fractions at individual CpGs. On all three platforms, cases are more highly methylated than controls across the DMR and at the adjacent boundary of the unmethylated region, where methylation rises abruptly in cases but gradually in controls. PacBio genomes from both groups show higher methylation than the other platforms within the unmethylated region and lower methylation in the flanking methylated regions, consistent with the platform offset in Figure 5, yet the case–control difference across the DMR is preserved. Ticks below panel C mark CpGs assayed by the methylation array.

PacBio genomes differed from the other platforms in one respect. For all three signatures, PacBio controls fell at the edge of the control cluster nearest the cases **(Figure 6A–C)**. For Kabuki syndrome the shift was large enough that one PacBio control, Control-11-PB, lies between the control and Kabuki clusters, nearest the Kabuki cases **(Figure 6C)**, and it was the only control called positive by the Kabuki classifier (probability 0.73; see below). The ONT (Control-11-ONT) and Illumina 5-base (Control-11-ILM) genomes from the same individual clustered with controls, as did Control-11-PB itself when the DMRs of all three signatures were analyzed together **(Figure 6E)**. Because this individual’s methylome resembles that of a control on two other platforms, we interpret the result as a platform-specific shift at the Kabuki DMRs to which the current Kabuki signature is not as robust, rather than as evidence of a Kabuki episignature in this individual. This matches the genome-wide comparison, in which PacBio differed most from the other two platforms **(Figure 5)**.

We then asked whether publicly available array data separate along the same axis at the Kabuki DMRs using our genome-derived classifier. Public 450K array data from individuals with *KMT2D* variants and controls (GSE97362; **Supplemental Table 5**)^22^ separated along the same principal component as the sequencing data **(Figure 6E)**, while the second principal component separated samples by platform within each group, consistent with the platform offset reported by Hildonen *et al*.^8^

Finally, we applied the final models to every library that was not used to train them, including PacBio and Illumina 5-base genomes from training individuals **(Figure 8)**. The *SETD1B*-NDD classifier assigned probabilities of 0.91–0.96 to the six held-out *SETD1B*-NDD ONT genomes (one from Erasmus MC and five from UBC; see below) and no more than 0.16 to any other library on any platform. The Kleefstra classifier assigned 0.70–0.97 to all eight held-out Kleefstra libraries (four ONT and four Illumina 5-base), and no other library exceeded approximately 0.51, although six fell in the indeterminate zone, four of them PacBio genomes. The Kabuki classifier assigned 0.97 or higher to five of six Kabuki libraries; the sixth, Kabuki-09-ILM, scored 0.55 and was flagged as indeterminate. Kabuki-09-ILM is an Illumina 5-base genome from Kabuki-09, an individual with a *KMT2D* missense variant whose DNA was extracted from a dried blood spot. The Kabuki classifier also made the most errors among libraries from other groups: Control-11-PB (0.73) and one PacBio Kleefstra genome (0.66) were called positive, and four others, three of them PacBio genomes, were indeterminate. Across the Kabuki and Kleefstra classifiers, 9 of the 12 indeterminate or false-positive calls among libraries from other groups were PacBio genomes, consistent with the PacBio offset described above.

**Figure 8.**
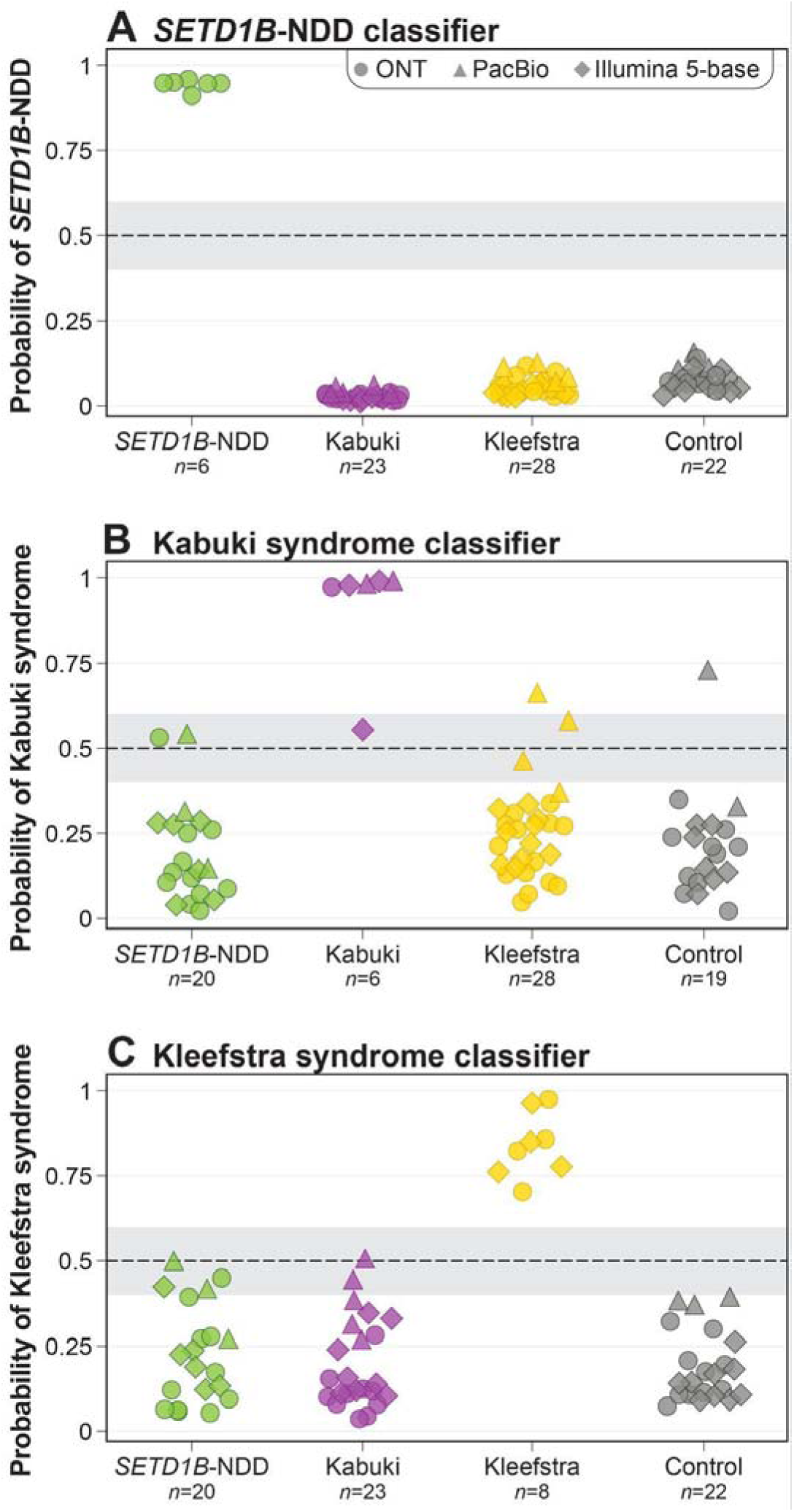
LRS-derived classifier scores for held-out sequencing libraries across platforms. Each panel shows the probability of disease assigned by one classifier to every library that was not used to train it. Panels show the **(A)** *SETD1B*, **(B)** Kabuki and **(C)** Kleefstra classifiers. Libraries are grouped by true diagnosis on the *x*-axis, colored by condition and shaped by platform. Libraries from individuals in the training set that were sequenced on a different platform are included, so one individual can contribute more than one point. The dashed line marks the 0.5 classification threshold, and the shaded band marks the indeterminate zone (0.4–0.6).

### Classification of genomes from outside sites

To test whether the classifiers hold up for samples collected and processed at other centers, we classified genomes from individuals contributed by two outside sites, none of whom contributed to DMR discovery or model training **(Table 1)**. The first site, UBC, evaluated five individuals with *SETD1B*-NDD, whose DNA was extracted from whole blood collected in EDTA tubes. The *SETD1B*-NDD classifier scored all five individuals above 0.9 **(Figure 8A)**, and the array-derived *SETD1B* classifier also called all five (0.40–0.74). These five genomes had higher coverage (29–46x) and longer reads (read N50 11.2–13.8 kb) than the *SETD1B*-NDD ONT genomes sequenced at UW in our study (18.2–25.0x and 3.9–11.4 kb), of which the array-derived classifier called only 4 of 11, so coverage or DNA quality may explain some of its misses.

The second site, UMC Utrecht, contributed 10 ONT genomes from nine individuals: three with Kleefstra syndrome, one of whom was sequenced twice; four with Kabuki syndrome, two with *KMT2D* and two with *KDM6A* variants; and two with *SETD1B*-NDD. These genomes differed from the UW samples in DNA extraction, fragmentation, library preparation, and basecalling model (HAC rather than SUP), and had a median coverage of 53.5x (12.2–78.6x) and a median read N50 of 9.9 kb (4.6–19.7 kb). The *SETD1B*-NDD classifier called both *SETD1B*-NDD cases, with probabilities of 0.96 and 0.77 **(Figure 9A)**, and assigned no more than 0.07 to any of the other eight genomes, including the lowest-coverage genome (12.2x). In principal component analysis, UMCU-025 grouped with the training cases, UMCU-244 lay between the two clusters but nearer the cases, consistent with its lower probability, and all eight other genomes fell on the control side of the first principal component **(Figure 9B)**. The same separation was visible at individual DMRs, where methylation in UMCU-025 resembled that of the training cases and methylation in UMCU-725, an individual with Kabuki syndrome, resembled that of the training controls **(Figure 9D, E)**. The array-derived classifiers also called most of these individuals correctly **(Figure 9C)**. The array-derived Kleefstra classifier called all three individuals with Kleefstra syndrome (0.87–3.16), and, as in our cohort, both genomes from one of them also scored above 0.3 for TBRS (0.54 and 0.79), though below their Kleefstra scores (3.16 and 2.85). The array-derived Kabuki classifier called all four individuals with Kabuki syndrome, including both with *KDM6A* variants, although their scores (0.44 and 0.61) were lower than those of the two with *KMT2D* variants (1.48 and 1.91). Finally, the array-derived *SETD1B* classifier again performed least well, calling one *SETD1B*-NDD case (0.74) and assigning the other to control (−0.76), the same case that the sequencing-derived classifier called with a probability of 0.77. The two genomes from the individual sequenced twice gave concordant results on every classifier **(Figure 9A–C)**.

**Figure 9.**
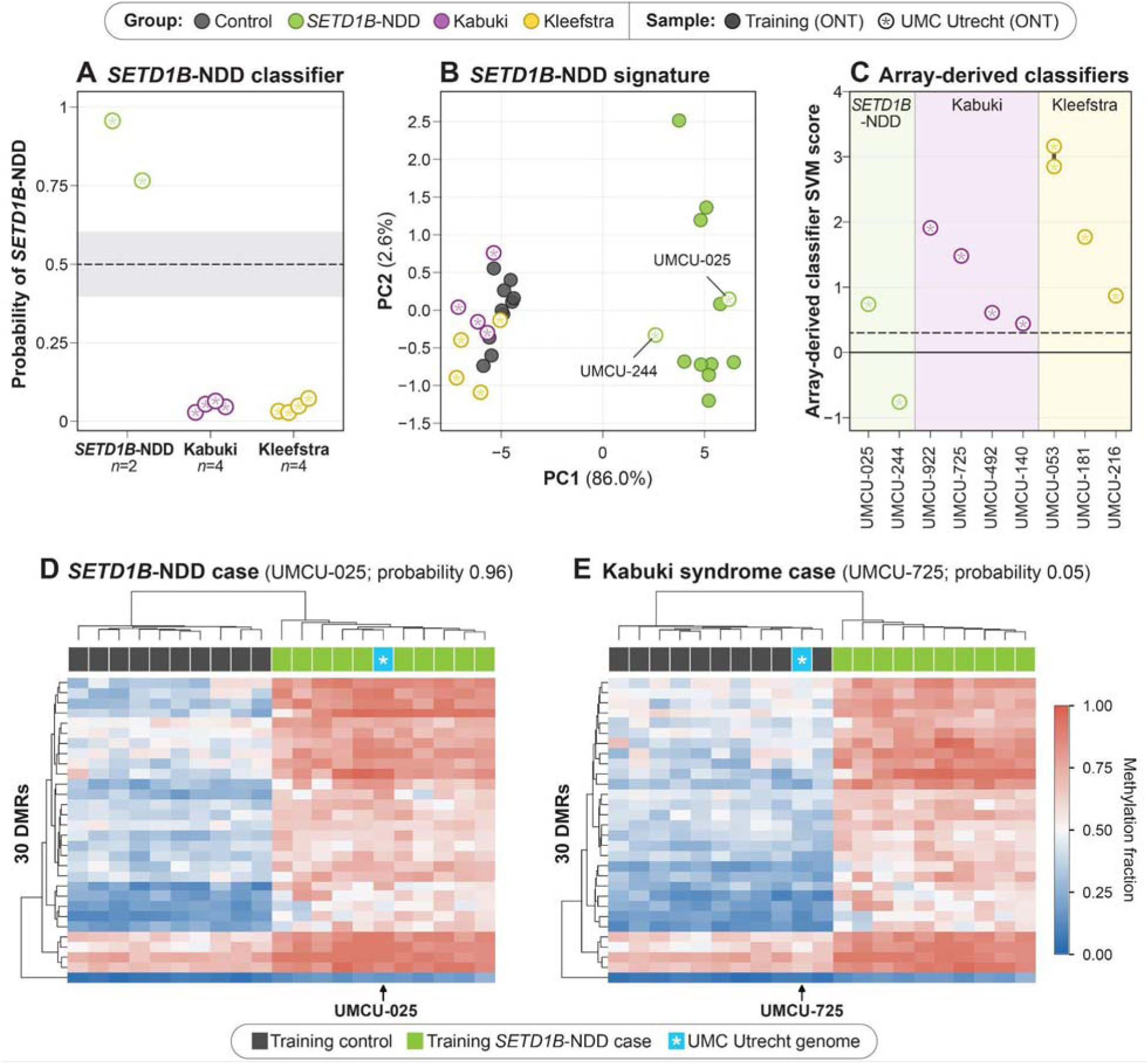
Classification of genomes sequenced at UMC Utrecht. All 10 genomes were classified with EpiSignaLR using an earlier release of the *SETD1B*-NDD model that used 30 DMRs rather than the 23 in the final model, so panels A, D, and E show these 30 DMRs; none were used for DMR discovery or model training. **(A)** *SETD1B*-NDD classifier probabilities for the UMC Utrecht genomes, grouped by diagnosis. **(B)** PCA of methylation at the DMRs selected by the *SETD1B*-NDD classifier. **(C)** Array-derived classifier scores for each UMC Utrecht individual on the classifier for that individual’s condition. **(D, E)** Methylation at the *SETD1B*-NDD classifier DMRs for the training cohort and one UMC Utrecht genome (*): **(D)** a *SETD1B*-NDD case classified as positive (UMCU-025, probability 0.96) and **(E)** a Kabuki syndrome case classified as negative (UMCU-725, probability 0.05).

### Sample collection method

Samples were collected in three ways, and the collection method was largely correlated with diagnosis **(Table 1)**. All *SETD1B*-NDD samples were venous blood, all Kleefstra syndrome samples were capillary blood collected with a Tasso device, Kabuki syndrome samples were capillary blood (*n* = 5), venous blood (*n* = 2), or dried blood spots (*n* = 5), and controls were capillary (*n* = 18) or venous (*n* = 5) blood. Collection method therefore could not be tested as an independent variable, but the data argue against a large effect. The four Kabuki cases in the training cohort whose DNA came from dried blood spots all scored as cases in cross-validation, and the array-derived Kabuki classifier correctly called all three individuals with *KMT2D* variants whose DNA came from dried blood spots **(Figure 2B)**. The array-derived Kabuki classifier missed both individuals in our cohort with *KDM6A* variants, whose DNA came from dried blood spots, but called both individuals with *KDM6A* variants sequenced at UMC Utrecht, whose DNA came from venous blood, although with lower scores than the two UMC Utrecht individuals with *KMT2D* variants. Because the UMC Utrecht samples also differed in DNA extraction, library preparation, and basecalling, and only four individuals with *KDM6A* variants were studied, we cannot tell whether sample type, these technical differences, or a weaker *KDM6A* signal at these CpGs explains the missed calls. Among ONT controls, the highest array-derived score did not differ significantly between capillary and venous samples (median −0.18 and −0.46; Mann-Whitney *p* = 0.10). Every PacBio and Illumina 5-base control was a capillary sample, so the collection method cannot be separated from the platform for the false-positive and indeterminate calls among controls on those platforms. The one held-out Kabuki genome flagged as indeterminate by the sequencing-derived classifier, Kabuki-09-ILM, also used DNA isolated from a dried blood spot **(Figure 8B)**; we consider this result in the Discussion.

## DISCUSSION

We have shown that published array-derived episignature classifiers transfer unevenly to genome sequencing data, and that episignatures can instead be derived directly from ONT methylation data and applied through a single workflow to genomes from other sequencing platforms. Array-derived classifiers performed well for Kleefstra syndrome but poorly for several individuals with *SETD1B*-NDD and for both individuals with Kabuki syndrome and *KDM6A* variants. For *SETD1B*-NDD, Kabuki syndrome, and Kleefstra syndrome, signatures developed from genome-wide methylation data ranked every case above every control in nested cross-validation, and roughly a quarter of the *SETD1B*-NDD and Kleefstra syndrome DMRs fall in regions with no array probe, demonstrating that key regions may have been overlooked by array-based analysis. Most Illumina 5-base and PacBio genomes clustered with ONT genomes from the same condition, and the final classifiers called every held-out case correctly except one indeterminate Kabuki genome. The *SETD1B*-NDD classifier also correctly called all seven *SETD1B*-NDD cases from two outside sites, UBC and UMC Utrecht, the latter of which used different DNA extraction, library preparation, and basecalling methods; as with several of the UW individuals, the array-derived classifier did not correctly classify one of these cases **(Figure 9)**.

As others have reported, we were able to apply published array-derived classifiers to our sequencing data, but found that their performance varied by condition **(Figure 2)**. The array-derived *SETD1B* classifier called only 5 of the 11 individuals sequenced at UW with *SETD1B*-NDD on any platform and none of the six Illumina 5-base genomes, and it called only one of the two cases sequenced at UMC Utrecht, whereas our sequencing-derived classifier called all eight held-out cases, including five from UBC and two from UMC Utrecht, with probabilities of 0.77 or higher. The array-derived Kabuki classifier missed both individuals with *KDM6A* variants in our cohort, whose DNA came from dried blood spots, and assigned a high score to one of them for Rubinstein-Taybi syndrome, but it called both individuals with *KDM6A* variants sequenced at UMC Utrecht from venous blood. Additionally, most individuals with Kleefstra syndrome scored above the threshold for Tatton-Brown-Rahman syndrome, although their Kleefstra score was always higher **(Figure 3)**. Two factors likely contribute to these discrepancies. First, methylation values from sequencing differ systematically from those measured on arrays, so thresholds set on array data may not hold for sequencing data.^7,8^ Second, the array-derived classifiers were trained on published median methylation values, one profile per condition, rather than on individual-level array data, so each one-versus-all classifier was built from a single positive example. A classifier trained on medians has no information about how much individuals vary at each CpG, so it cannot give less weight to variable CpGs, set its threshold from the spread of real controls, or be recalibrated for sequencing data. Individual-level data would therefore likely improve calibration and specificity **(Figure 3)**. Because the data underlying these medians are not publicly available, we could not determine whether these off-target calls reflect overlap between signatures, a platform effect, or a characteristic of these individuals. For clinical testing, this means that a negative or ambiguous result from an array-derived classifier applied to sequencing data may not be able to be taken at face value, that performance must be established for each condition and platform, and that conditions such as *SETD1B*-NDD, for which the array signature transfers poorly, may need to be recreated using data generated by sequencing to be trustworthy.

Kabuki syndrome was the condition for which our classifier made the most errors, and it is also the only one of the three with a recently refined array signature against which our results can be compared, the KS1 episignature reported by Wang and colleagues.^5^ That signature was discovered using EPIC v1.0 arrays in 25 individuals with *KMT2D* loss-of-function variants and 43 controls, contains 589 CpGs, and refines an earlier signature discovered on 450K arrays in 11 individuals. Their strongest changes were in the HOXA cluster, where we independently found a hypomethylated DMR spanning *HOXA3* and *HOXA4*. Their signature also classified individuals with pathogenic *KDM6A* variants as KS1-like, which supports our inclusion of two such individuals in our training set. Although the refined and original signatures shared only 78 CpGs (13%), both classified most samples correctly, which mirrors the even smaller overlap between our sequencing-derived signature and the original array-derived Kabuki classifier (2 of 153 CpGs). Unlike ours, both of their signatures were discovered on arrays, and PacBio genomes were used only for testing, using methylation values at array probe positions. On these genomes, the original signature classified five of eight controls as intermediate, whereas the refined signature classified all eight as controls.^5,22^ Our Kabuki signature, built with 13 DMRs from 19 genomes, is at a similar stage to the original signature, and their experience suggests that a larger and more diverse training cohort will improve performance across platforms more than focusing on a single platform will.

Only one held-out case, Kabuki-09-ILM, was not confidently classified, with a Kabuki probability of 0.55 **(Figures 6C and 8B)**. This individual carries a rare, likely damaging *KMT2D* missense variant rather than the truncating variants carried by most of our cohort, supported by Wang and colleagues finding that some pathogenic *KMT2D* missense variants produce intermediate scores on their classifier.^5^ The array-derived Kabuki classifier called the same library positive **(Figure 2B)**, which argues against a technical failure of the dried blood spot DNA or the library and suggests that this variant changes methylation less at our 13 DMRs than at the array CpGs. Two training cases with *KMT2D* missense variants scored as cases, so the effect is likely specific to this variant. Mosaicism is another possibility, as Wang and colleagues found that a mosaic *KMT2D* frameshift variant, detected by PacBio sequencing, explained a lower classifier score in one of their cases. Until signatures are trained on samples that are unique to individual genes, span variant types, collection methods, platforms, and ages, an intermediate result should prompt confirmatory testing rather than be read as negative.

Generally, data generated on the PacBio platform behaved differently from the ONT and 5-base data. One PacBio control, Control-11-PB, was called positive by the Kabuki classifier even though the ONT and Illumina 5-base genomes from the same individual were grouped with controls **(Figure 6C)**. PacBio controls were also shifted toward cases for every signature, PacBio reported many fully methylated CpGs as partially methylated **(Figure 5)**, and PacBio genomes accounted for most indeterminate and false-positive calls **(Figure 8)**. This could reflect how each platform calls modified bases, how CpG calls are summarized, or how coverage varies at individual DMRs, but without a large number of reference methylomes from unaffected individuals generated on each platform we cannot determine which factor is the largest driver of these differences. Early array-based episignature studies shared data that others could build on, yet today only EpigenCentral allows others to apply and evaluate published classifiers.^4,5^ This demonstrates how sequencing-based episignatures will benefit from broad sharing of methylation data, importantly to include platform-matched control methylomes, consented case data, and code and models that can be retrained as data accumulate. For this reason, we have made the methylation data, signatures, and code from this study publicly available and plan to do so going forward.

This study has several limitations. Each signature was built from nine or 10 cases and 10 controls, which left the Kabuki probability scale poorly calibrated and the *SETD1B*-NDD panel unstable, and cases and controls also differed in age, collection method, and, for Illumina 5-base, sequencing depth. Validation at outside sites was also limited: the seven *SETD1B*-NDD cases from UBC and UMC Utrecht did not include unaffected controls from those sites, so the specificity of the *SETD1B*-NDD classifier there was assessed only against individuals with Kabuki or Kleefstra syndrome sequenced at UMC Utrecht. Addressing these limitations will require larger, age-matched cohorts sequenced on each platform, and because these conditions are individually rare, no single center can assemble such cohorts alone. This is where the field has struggled. The individual-level data behind many published episignatures, including the array-derived classifiers we applied here, are not publicly available, so these classifiers cannot be independently evaluated, recalibrated for new platforms, or extended as new cases are identified. Methylation is now a native output of ONT, PacBio, and Illumina 5-base genomes, so for an individual already undergoing genome sequencing, episignature analysis adds mainly computational cost, and the main barrier to its clinical use is access to data rather than technology. We are therefore committed to openly sharing the methylation data, signatures, and code from this study and from our future work so that others can test, retrain, and build on them. Sharing of this kind will allow sequencing-based episignatures to become a resource for the entire rare disease community, advancing our understanding and diagnosis of rare genetic conditions.

## DATA AND CODE AVAILABILITY

The EpiSignaLR workflow can be found on GitHub: https://github.com/millerlaboratory/EpiSignaLR. This includes the training data for the three ONT-derived signatures. Code used in this study can be found on GitHub: https://github.com/millerlaboratory/EpiSignaLR

## USE OF AI-ASSISTED TOOLS

During preparation of this manuscript, the authors used Claude (Anthropic, Opus 5.5) to edit text for clarity and consistency, draft and revise figure legends, check numerical statements in the text against the supplemental tables, check references, and generate plotting code used to redraw and lay out figures. AI tools were not used to generate, modify or analyze sequencing data, to produce genotype calls, or to make analytic or interpretive decisions. All AI-assisted content was reviewed, verified and edited by the authors, who take full responsibility for the content of the publication.

## ACKNOWLEDGEMENTS

We thank Combined Brain and the families who contributed samples to its biorepository. We thank Geoffroy Delplancq (CH Versailles, Le Chesnay, France) for his contributions to this study. The Barakat lab was supported by the Netherlands Organisation for Scientific Research (ZonMw Vidi, grant 09150172110002), and acknowledges ongoing support for rare disease research from Stichting 12q. Funding bodies did not have any influence on study design, results, and data interpretation or final manuscript. WTG’s research salary is supported by the BC Children’s Hospital Foundation through an intramural IGAP award. WTG’s work was supported by the BCCHRI’s Canucks for Kids Fund and by CIHR Project Grant PJT-168982. SJMJ is funded by the Canada Research Chair program and by CIHR Project Grant PJT-185999. DEM is supported by NIH grant DP5OD033357. We acknowledge referring clinicians Dr. Elizabeth Digby and Dr. Cyrus Boelman at BC Children’s Hospital.

## AUTHOR CONTRIBUTIONS

Conceptualization: DEM

Methodology and software: SBG, DEM, MPGZ

Investigation: SBG, MPGZ, JG, EJH, RL, JRR, SHRS, KLe, MGE, TP, ZBA, DM

Resources: TSB, YuS, KLa, WTG, YaS, SJMJ, SV, MHW, JRR, KLIvG

Visualization: SBG, ALM, DEM

Writing, original draft: SBG, DEM

Writing, review and editing: all authors

## REFERENCES

1. Aref-Eshghi, E. et al. Evaluation of DNA Methylation Episignatures for Diagnosis and Phenotype Correlations in 42 Mendelian Neurodevelopmental Disorders. Am J Hum Genet 106, 356–370 (2020).

2. Sadikovic, B. et al. Clinical epigenomics: genome-wide DNA methylation analysis for the diagnosis of Mendelian disorders. Genet. Med. 23, 1065–1074 (2021).

3. Levy, M. A. et al. Novel diagnostic DNA methylation episignatures expand and refine the epigenetic landscapes of Mendelian disorders. HGG Adv 3, 100075 (2022).

4. Turinsky, A. L. et al. EpigenCentral: Portal for DNA methylation data analysis and classification in rare diseases. Hum. Mutat. 41, 1722–1733 (2020).

5. Wang, M. et al. A next-generation episignature for Kabuki syndrome enables fine mapping of the impact of KMT2D variants to inform precision medicine. Am. J. Hum. Genet. 113, 1233–1252 (2026).

6. Illumina 5-Base DNA Prep Product Documentation. https://support.illumina.com/content/dam/illumina-support/documents/documentation/chemistry_documentation/illumina-5-base-dna-prep/200073497_01_illumina-5-base-dna-prep-prod-doc.pdf (2026).

7. Geysens, M. et al. Clinical evaluation of long-read sequencing-based episignature detection in developmental disorders. Genome Med. 17, 1 (2025).

8. Hildonen, M. et al. Clinical feasibility of long-read WGS for DNA methylation signature analysis. Clin. Genet. 109, 725–729 (2026).

9. Li, H. Minimap2: pairwise alignment for nucleotide sequences. Bioinformatics 34, 3094–3100 (2018).

10. Zheng, Z. et al. Symphonizing pileup and full-alignment for deep learning-based long-read variant calling. Nature Computational Science 2, 797–803 (2022).

11. Smolka, M. et al. Detection of mosaic and population-level structural variants with Sniffles2. Nat. Biotechnol. 1–10 (2024).

12. Lin, J.-H., Chen, L.-C., Yu, S.-C. & Huang, Y.-T. LongPhase: an ultra-fast chromosome-scale phasing algorithm for small and large variants. Bioinformatics 10.1093/bioinformatics/btac058 (2022) doi:10.1093/bioinformatics/btac058.

13. PacBio. *Pb-CpG-Tools*. (Github).

14. Illumina. DRAGEN secondary analysis (Version 4.5) [Computer software]. https://www.illumina.com/products/by-type/informatics-products/dragen-secondary-analysis.html (2026).

15. Oxford Nanopore Technologies. Modkit: A Bioinformatics Tool for Working with Modified Bases. (Github).

16. Korthauer, K., Chakraborty, S., Benjamini, Y. & Irizarry, R. A. Detection and accurate false discovery rate control of differentially methylated regions from whole genome bisulfite sequencing. Biostatistics 20, 367–383 (2019).

17. Hansen, K. D., Langmead, B. & Irizarry, R. A. BSmooth: from whole genome bisulfite sequencing reads to differentially methylated regions. Genome Biol. 13, R83 (2012).

18. Zou, H. & Hastie, T. Regularization and variable selection via the elastic net. J. R. Stat. Soc. Series B Stat. Methodol. 67, 301–320 (2005).

19. Platt, J. C. Probabilistic Outputs for Support Vector Machines and Comparisons to Regularized Likelihood Methods. in Advances in Large Margin Classifiers 61–74 (MIT Press, 1999).

20. VaroquauxGaël & DuchesnayÉdouard. Scikit-learn: Machine Learning in Python. J. Mach. Learn. Res. 10.5555/1953048.2078195 (2011) doi:10.5555/1953048.2078195.

21. Quinlan, A. R. & Hall, I. M. BEDTools: a flexible suite of utilities for comparing genomic features. Bioinformatics 26, 841–842 (2010).

22. Butcher, D. T. et al. CHARGE and Kabuki syndromes: Gene-specific DNA methylation signatures identify epigenetic mechanisms linking these clinically overlapping conditions. Am. J. Hum. Genet. 100, 773–788 (2017).

